# ShaPRS: Leveraging shared genetic effects across traits or ancestries improves accuracy of polygenic scores

**DOI:** 10.1101/2021.12.10.21267272

**Authors:** M. Kelemen, E. Vigorito, L. Fachal, C. A. Anderson, C. Wallace

**Author notes:** These authors contributed equally (C. A. Anderson, C. Wallace).

## Abstract

We present shaPRS, a novel method that leverages widespread pleiotropy between traits, or shared genetic effects across ancestries, to improve the accuracy of polygenic scores. The method uses genome-wide summary statistics from two diseases or ancestries to improve the genetic effect estimate and standard error at SNPs where there is homogeneity of effect between the two datasets. When there is significant evidence of heterogeneity, the genetic effect from the disease or population closest to the target population is maintained. We show via simulation and a series of real-world examples that shaPRS substantially enhances the accuracy of PRS for complex diseases and greatly improves PRS performance across ancestries. shaPRS is a PRS pre-processing method that is agnostic to the actual PRS generation method and, as a result, it can be integrated into existing PRS generation pipelines and continue to be applied as more performant PRS methods are developed over time.

## Introduction

Genome-wide association studies (GWAS) provide a routine means of quantifying the effects of genetic variation on human diseases and traits. One possible use of these genetic effect estimates is the creation of polygenic risk scores (PRSs), an approximation of an individual’s genome-wide genetic propensity for a given trait or disease. Recent studies have shown that individuals in the upper extreme tail of polygenic risk for some common diseases have equivalent risk to those carrying monogenic mutations for these phenotypes^1,2^. Driven by these observations there is hope that polygenic scores can be used alongside traditional clinical and demographic predictors of disease to diagnose disease earlier and with greater accuracy^3,4^.

Unfortunately, the clinical utility of polygenic scores is currently limited by the GWAS on which they are based. The precision with which GWAS can estimate genetic effects on disease risk increases with sample size. Recent studies have suggested that most complex diseases will require somewhere between a few hundred thousand to several million cases to accurately capture genome-wide genetic effects on disease risk^5,6^. As a result, the information content of all current GWAS is imperfect, reducing the accuracy of the polygenic scores generated from them. There is an expectation that GWAS meta-analyses across vast population biobanks will get us closer to quantifying SNP effects that fully capture heritability for some common complex diseases. However, many debilitating and life-threatening complex diseases have lower population prevalence, preventing even meta-analyses across large biobanks from ascertaining sufficient cases to facilitate the construction of accurate polygenic scores.

It is not only less common complex diseases that are set to be precluded from any clinical advantages brought about by polygenic scores. Genomics is failing on diversity^7^. On February 14th, 2024 the GWAS Diversity Monitor^8^ showed that 94.51% of individuals included in GWAS were from European ancestries. Recent studies have demonstrated the poor portability of polygenic risk scores across populations due to differences in effect sizes and LD structure^9^. Migration events and population bottlenecks can lead to large differences in allele frequencies between ancestries and, as a result of the biased application of GWAS, we are missing accurate disease risk estimates for the many variants that are only common outside of European ancestry groups^10,11^. Thankfully, the clarion call for major improvements in the ancestral diversity of GWAS, and genomics studies more generally, is now loud^7,12,13^. Recent studies in non-Europeans have highlighted the advantages of increased diversity of GWAS, delivering both novel genetic associations and biological insights that were missed even in the larger European ancestry GWAS studies^9,14–16^. If polygenic risk scores do start to deliver on their hype then further diversification cannot come soon enough – otherwise we run the risk of widening existing health inequalities.

While genetic effects on disease certainly do differ between populations, many risk variants are believed to be shared across divergent ancestry groups^17,18^. There is also a growing appreciation of the extent to which genetic effects are shared across different disorders. For clinically and biologically related diseases such as Crohn’s disease and ulcerative colitis, the two common forms of inflammatory bowel disease, genetic effects are often shared. Across immune-mediated disease more generally the number of known pleiotropic effects continues to grow, a phenomenon that is mirrored in other disease groups such as metabolic and psychiatric disorders. A principled pooling of information across traits^19,20^ and ancestries^21–23^ has already been shown to improve prediction accuracy of PRS. A common assumption of these methods is that weights given to each dataset are constant across SNPs. In reality, this assumption is frequently violated as the extent of sharing, either between two diseases or two populations, varies across SNPs^24,25^.

We introduce a novel method, shaPRS (pronounced Shapers), a PRS pre-processing step that can be integrated into existing PRS generation pipelines that allows integration of imperfectly shared information between two GWAS datasets. We assume one dataset is representative of the target population, hereafter referred to as the proximal dataset, and that a second adjunct dataset may provide relevant information but that the degree of relevance varies across the genome. Our approach, which only requires summary statistics for each dataset, estimates weights that summarise how relevant the adjunct dataset is at each SNP to perform a weighted meta-analysis of the two datasets. We also generate a new pairwise SNP correlation matrix that captures the effect of this weighting, and allows for partial sharing of controls and/or the use of distinct SNP correlation matrices for the two input studies (eg in the case of different ancestries). This matrix may be used together with the weighted SNP effect estimates in any downstream PRS software. We show in large-scale simulations in the UK Biobank (UKBB)^26^ that shaPRS outperforms similar methods. We then apply shaPRS to nine real GWAS datasets to illustrate the improvements it brings to PRS accuracy, both across diseases and across ancestral populations.

### Overview of method

shaPRS, which uses GWAS summary statistics, is a PRS pre-processing step based on a weighted meta-analysis of two partially related GWAS studies. We begin by testing, at each SNP, evidence against homogeneity of effect between the two studies using Cochran’s test. From these test statistics, we calculate the local false discovery rate (lFDR)^27^ as an estimate of the probability that the estimates reflect the same “common truth”. Where the lFDR is high, it is likely that the datasets can be combined and we favour *β*_12_, which is the standard inverse variance weighted average of the effect estimates in the proximal study, *β*_1_, and the adjunct study, *β*_2_. Our aim is to minimise variance of estimated effect sizes by including information from the adjunct study, where doing so is unlikely to cause bias. Where the lFDR is low, we are conservative, and favour *β*_1_ from the proximal study, aiming to minimise bias at the expense of higher variance. We thus calculate a final shaPRS SNP effect estimate as

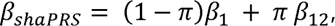

where *π* denotes the lFDR. As the use-case of our method is a seamless integration into existing PRS generation pipelines, a full set of summary statistics are derived, including standard errors, p-values and sample size, as described in the Materials and Methods section. An illustrative example is provided in the Supplementary Information.

The current generation of most performant PRS generation methods^28–30^ also require an appropriate LD-matrix, often obtained from a reference panel. Therefore, to obtain an LD-reference panel appropriate for the derived summary statistics that represent information from the weighting and possibly LD from different ancestries, we provide a method to derive a new matrix describing the correlation between *β_shaPRS_* across different SNPs (Supplementary Note).

## Materials and methods

### ShaPRS genetic association summary statistics blending

Our approach is based on a weighted averaging of each SNP’s estimated effect between a single proximal dataset and an inverse variance meta-analysis of the proximal and adjunct datasets. The full derivations are set out in the Supplementary Note, and summarised here. Our method favours the proximal dataset effect estimate *β_1_* where the effect estimates appear to differ between proximal and adjunct datasets, and the combined effect estimate *β_12_* (the standard fixed effects meta-analysis estimate obtained from *β_1_* and the adjunct study coefficient *β_2_*) when the effect estimates for the two datasets are similar. In other words, we choose the more precise proximal phenotype with lower bias where SNP effects are heterogeneous, but prefer the larger sample size with lower variance where the SNP effects are congruent between single datasets.

To make this decision, we use Cochran’s Q-test to assess heterogeneity of effects between the two datasets at each variant, modified to allow for shared controls between the cohorts.

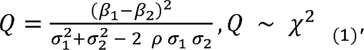

where □*_1_* / □*_2_* are the standard errors for the proximal and adjunct datasets, respectively, and *ρ* is an estimate of the correlation between *β_1_* and *β_2_* obtained as a simple function of sample sizes^31^.

To estimate the probability that effects are heterogeneous, we used a local FDR approach, estimating

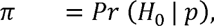

where *H_0_* is the null hypothesis for the SNP, and *p* is the (adjusted) Q-test p-value obtained from the Chi-squared distribution with one degree of freedom as defined above. The lFDR values were then estimated from these p-values by the *qvalue* R package^32^.

The blended effect estimate is then

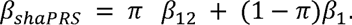

The goal of our method is to generate a new, complete set of summary statistics that may be used by any downstream PRS generation tool. These statistics include a new set of SNP coefficients, their standard errors and the correlation between coefficients. The Supplementary Note sets out derivations for the standard errors and correlation matrix, and functions to calculate these are provided in the R package https://github.com/mkelcb/shaprs.

### Simulation analyses

Our simulations relied on the UK Biobank (UKBB) cohort, which has been previously described in detail elsewhere^26^. We excluded individuals who were sex-discordant, not ‘white British’ or had third-degree relatives or closer in the cohort, as defined in the UK Biobank documentation. Genotype data were filtered to an intersection of the HapMap3 panel (a common practice for PRS generation, but not a requirement for shaPRS) and a subset that excluded variants with an INFO score <0.8, MAF <0.1%, missing genotype rate >2% or deviated from Hardy-Weinberg equilibrium (P<10^-7^). From this subset, we randomly chose 3,158 individuals to serve as a test set (approximately 20% of the size of our IBD dataset).

We evaluated the effect of cohort size by considering three studies with approximately half (N=7,022), equal to (N=14,044) or double (N=28,088) the number of samples in our genotyped IBD cohort, after withholding 3,158 individuals as a test set that were not used for model training. We also considered three different ratios to split our source samples into the two phenotypes (proximal and adjunct). These ratios were 20/80, 40/60 and 50/50 for phenotype 1 and 2, respectively. Additionally, we varied the range of pleiotropic architectures considered by evaluating three genetic correlations (0.1, 0.25 and 0.5) made up from three combinations of shared and non-shared SNP effects. The motivation for the latter was to examine the key ability of our method to adapt to different compositions of shared and non-shared genetic effects that comprise a given level of genetic correlation. We considered three different scenarios (low, medium and high, as defined in Table S1) of shared effects per genetic correlation, making up a total of nine arrangements. We also considered an additional scenario, where five SNPs contribute 5% of the total non-shared heritability for each trait. We evaluated all possible parameter combinations at a heritability of 0.5 arising from 1,000 causal variants for a set of 162 genetic architecture scenarios. We also evaluated the performance of all methods in the four additional scenarios where we have held all parameters at their base value except for one (the number of causal SNPs of 3,000 or 5,000, and a heritability of 0.25 or 0.75). The results of these additional 36 simulation scenarios can be found Fig S4. We used LDAK 5.0^33^ to simulate 20 replicates for bivariate quantitative phenotypes whose SNP effect sizes we generated via our custom R scripts according to the schema described above. We evaluated shaPRS’s performance by comparing its predictive accuracy on the test set against four baselines: the single proximal dataset on its own, the meta-analysis of the proximal and adjunct datasets and the SMTPred and MTAG methods. Both SMTPred and MTAG were trained directly on the PLINK summary statistics using their own python functions *’ldsc_wrapper.py’* and *’mtag.py’* for SMTPred and MTAG, respectively. To accommodate the scale of our simulations, the final PRS were generated via RapidoPGS, a computationally efficient PRS generation method^34^. To evaluate if using RapidoPGS had introduced any bias into our analyses, we re-generated the PRS of 50 randomly selected replicates (10 for each method) with LDpred2-auto. For this, we chose the scenario involving 14,044 individuals, phenotypes divided 50/50, with an rG of 0.5 made up from half of the causal variants shared with a correlation of 1.0, without any highly penetrant variants. We found the relative performance of the methods did not change and that the results were strongly congruent between LDpred2 and RapidoPGS (Spearman rank correlation of 0.781).

### Generating genome-wide summary statistics for Crohn’s disease and ulcerative colitis

The availability of all IBD datasets are described in the *Data and code availability* section. The sample collection protocols are described in the original publications of each study^35–37^. Initial quality control procedures for the studies where the IBD PRS performance was tested are described in the original publications^26,36,37^. For the IBD training dataset, and prior to genotype imputation, we excluded (1) A/T and C/G genotyped variants with MAF ≥0.45 in 1000GP EUR subset^38^; (2) variants with a call rate < 0.95 (or 0.98 call rate for variants with MAF < 0.01); (3) variants with a significant difference in genotype call rate between cases and controls (p-value <1x10^-4^); (4) variants with allele frequency differences versus those reported in Gnomad non-finish Europeans or TOPMed^39^ global MAF (using the criterion ((p1 − p0)^2/((p1+p0)*(2-p1-p0)) > 0.025 and >.125, respectively), where p0 is the MAF in the reference panel and p1 the observed MAF in the study); (5) variants with a HWE p-value < 10^-5^ among controls and 10^-12^ among cases; and (6) monomorphic variants. We also excluded samples with a missing genotype rate >0.5; a heterozygosity estimate +/- 4 standard deviations from the mean (per continental population), a mismatch between recorded gender and inferred genotypic sex, with a kinship coefficient ≥ 0.345 (defined using KING^40^ (v2.2.4)) with another sample within the study, or ≥0.177 with another sample in other UK IBD study; or with evidence of non-European ancestry, defined by projecting the samples onto principal components estimated from 1000 genomes project reference samples^38^.

The datasets were imputed using the multi-ancestry TOPMed reference panel (r2@1.0.0) via the TOPMed imputation server^39,41^ (imputationserver@1.5.7). After the first round of imputation, variants with an empirical R^2^ < 0.5 were excluded from analysis. Imputation was repeated after correcting strand issues at SNPs with an empirical R^2^ < -0.5. Post imputation, variants with HWE p-value ≤ 1x10^-5^, MAF < 0.001, or imputation R^2^< 0.4 were excluded. The GWAS training datasets included 4,647 and 5,400 UC and CD cases, respectively, and 10,308 shared controls.

Association tests for UC and CD were performed using Regenie^42^ (v1.0), including European populations principal components and sex as covariates. This produced GWAS summary statistics for 14,056,620 variants.

### Building polygenic risk scores from Crohn’s disease and ulcerative colitis GWAS summary statistics

Summary statistics for CD and UC were initially filtered to remove those SNPs with an imputation INFO (or MARCH R^2^) < 0.8 or those failing the following quality-control thresholds:

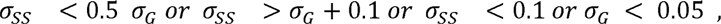

where σ*_ss_* is defined as

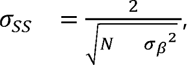

and σ _G_ as

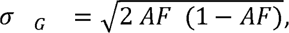

where AF, N and σ_*β*_^2^ are the minor allele frequency, the sample size and standard error of the SNP coefficients, respectively. The above threshold criteria used for this filtering step were sourced from the LDpred2 recommended settings. We then applied shaPRS across the remaining set of SNPs, which were then filtered to only keep SNPs that were either in the HapMap3 panel (a common practice for PRS generation) or had a trait heterogeneity lFDR < 1. We chose to expand beyond HapMap3 in the case of IBD, as many of the variants that differentiated CD from UC were not captured on the standard HapMap3 panel. However, we note that shaPRS is completely agnostic to the set of SNPs it is applied to, as it can be applied to any summary data from genome-wide SNPs to the typical HapMap3-based PRS panel. To accommodate the non-HapMap3 SNPs in the LD-reference panel, we generated SNP-SNP correlation matrices for 1,703 LD blocks^43^ using PLINK’s ‘ *--r*’ function. The resulting files were then compressed and packaged via a custom python script into the same *hdf5* format as used by PRS-CS. This procedure left 856,877 SNPs that were used to generate the PRS by all the evaluated methods. The final PRSs for the IBD datasets were built using PRS-CS and the profile scores for our test set individuals were generated using PLINK’s *’--score’* function.

### Cross-ancestry datasets and PRS model evaluation

The Japanese association summary data for the five traits (asthma, height, BRCA, CAD and T2D) were all retrieved from the Biobank Japan repository^44,45^. The European association data for the same five traits were sourced from different studies identified through the GWAS catalogue selected based on the criteria that they were of comparable sample size, and that they did not overlap with the (non-interim) UK Biobank (UKBB) release. For the scenario involving the African proximal population, we obtained African ancestry GWAS summary statistics from the Uganda Genome Resource^46^ for three traits (BMI, height and LDL cholesterol levels), with European adjunct association summary statistics from other published sources^47–49^. The full set of sources are shown in Table 1.

**Table 1.**
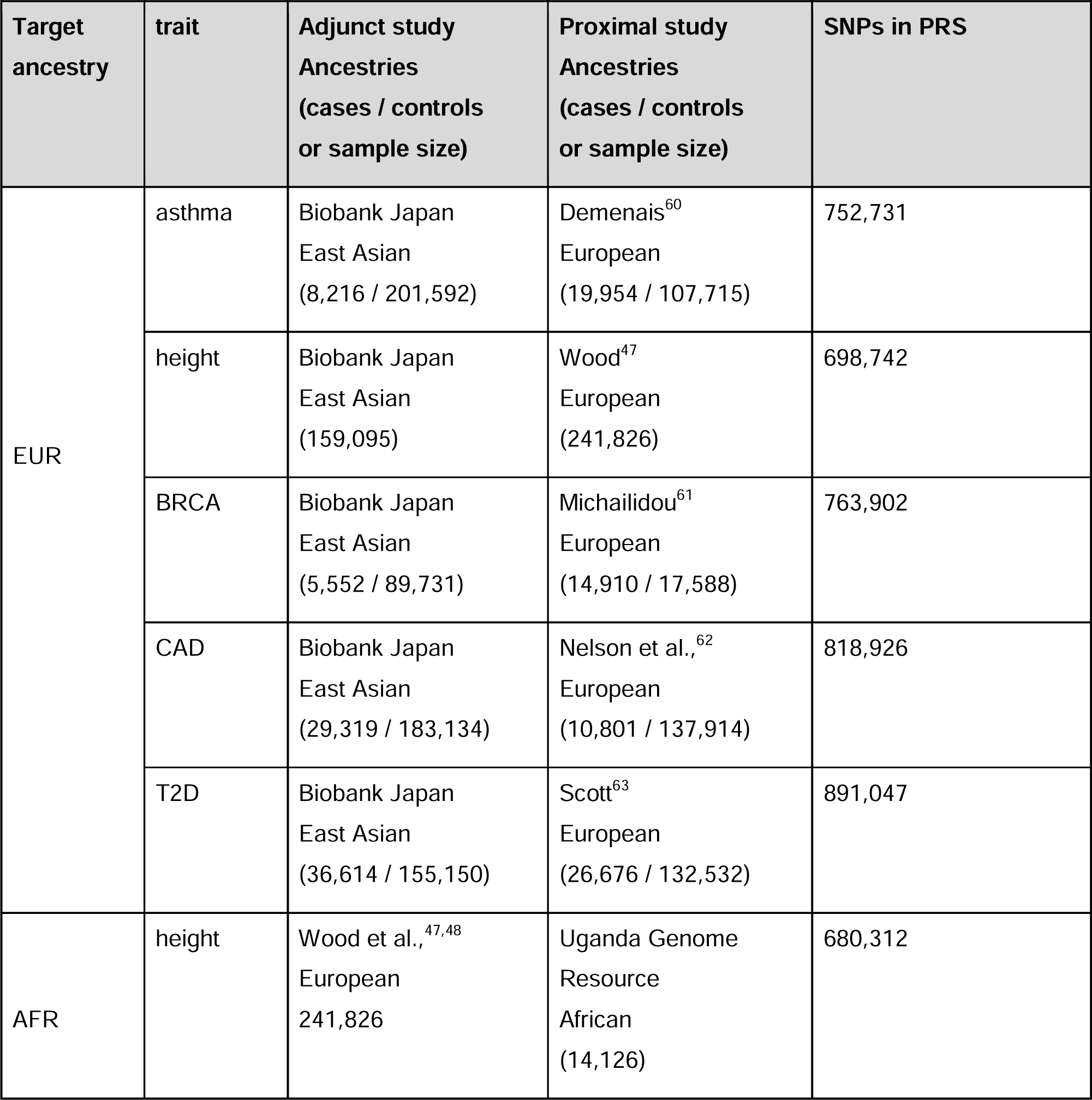
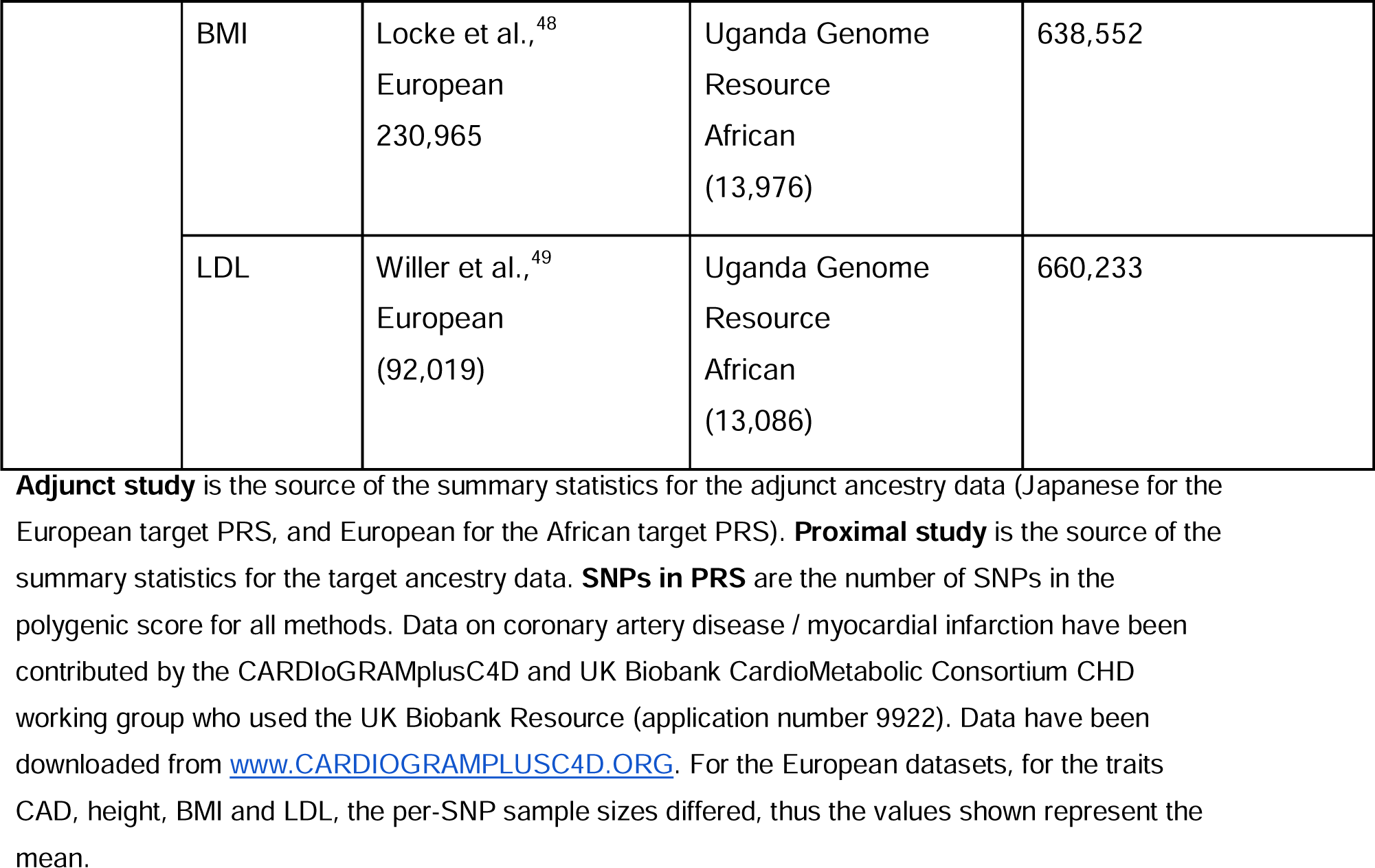
Cross-ancestry PRS data parameters.

| Target ancestry | trait | Adjunct study Ancestries (cases / controls or sample size) | Proximal study Ancestries (cases / controls or sample size) | SNPs in PRS |
| --- | --- | --- | --- | --- |
| EUR | asthma | Biobank Japan<br>East Asian<br>(8,216 / 201,592) | Demenais<sup>60</sup><br>European<br>(19,954 / 107,715) | 752,731 |
|  | height | Biobank Japan<br>East Asian<br>(159,095) | Wood<sup>47</sup><br>European<br>(241,826) | 698,742 |
|  | BRCA | Biobank Japan<br>East Asian<br>(5,552 / 89,731) | Michailidou<sup>61</sup><br>European<br>(14,910 / 17,588) | 763,902 |
|  | CAD | Biobank Japan<br>East Asian<br>(29,319 / 183,134) | Nelson et al.,<sup>62</sup><br>European<br>(10,801 / 137,914) | 818,926 |
|  | T2D | Biobank Japan<br>East Asian<br>(36,614 / 155,150) | Scott<sup>63</sup><br>European<br>(26,676 / 132,532) | 891,047 |
| AFR | height | Wood et al.,<sup>47,48</sup><br>European<br>241,826 | Uganda Genome<br>Resource<br>African<br>(14,126) | 680,312 |
|  | BMI | Locke et al.,<sup>48</sup><br>European<br>230,965 | Uganda Genome<br>Resource<br>African<br>(13,976) | 638,552 |
|  | LDL | Willer et al.,<sup>49</sup><br>European<br>(92,019) | Uganda Genome<br>Resource<br>African<br>(13,086) | 660,233 |

**Adjunct study** is the source of the summary statistics for the adjunct ancestry data (Japanese for the European target PRS, and European for the African target PRS). **Proximal study** is the source of the summary statistics for the target ancestry data. **SNPs in PRS** are the number of SNPs in the polygenic score for all methods. Data on coronary artery disease / myocardial infarction have been contributed by the CARDIoGRAMplusC4D and UK Biobank CardioMetabolic Consortium CHD working group who used the UK Biobank Resource (application number 9922). Data have been downloaded from www.CARDIOGRAMPLUSC4D.ORG. For the European datasets, for the traits CAD, height, BMI and LDL, the per-SNP sample sizes differed, thus the values shown represent the mean.

To produce the final LD-reference data, we used a custom R script (*’LDRefGen_wrapper.R’*, included in the project github) to estimate the pairwise correlations between shaPRS SNP effect estimates. This functionality is now available in our R package via the *shaPRS_LDGen* function, which requires the proximal and adjunct LD-reference panels in LDpred2 format and the shaPRS pre-processed summary data. For more information on the mathematical details on the LD derivation see the Supplementary Note. To maximise the fraction of variants available across ancestries and summary datasets, HapMap3 SNPs were chosen that were shared between the adjunct and proximal summary statistics that were also present in the UKBB imputed dataset with an INFO score > 0.8. The final PRS were built after the removal of ambiguous alleles (A/T and G/C). All PRS generation methods, LDpred2, PRS-CS and PRS-CSx, were applied via their respective ‘auto’ options to estimate overall shrinkage, keeping with our use-case of no additional genotype data being available to fine tune hyper-parameters. PRS profiles were generated in PLINK^50^ and evaluated using individual genotypes from the UK Biobank cohort. For all traits we excluded related individuals and restricted the analysis to individuals of the proximal ancestry. For the generation of the proximal African population we identified 6,414 individuals using the population centroids published by Prive et al^51^, and for the European adjunct dataset we relied on the flag “white British” ethnicity (UKBB field 21000, code 1001) in the UKBB documentation^51^. We also excluded ∼30,000 individuals from the initial release that were genotyped with the UK BiLEVE array. We identified those individuals using field “22000” batches coded -1 to -11. For BRCA, CAD and T2D we applied the same selection criteria for cases and controls as previously described^52^, using the same UKBB codes for each of the relevant traits as in https://github.com/privefl/simus-PRS/tree/master/paper3-SCT/code_real). Briefly, we included as cases those individuals who self-reported the condition or were diagnosed by a medical doctor or the condition was included in their death record. For breast cancer we excluded individuals with other cancer diagnosis and restricted the analysis to females (108,21 cases, 147.134 controls). For T2D we excluded individuals with type 1 diabetes (12,288 cases, 301,822 controls) and for CAD we excluded individuals with other heart conditions (10,611 cases, 209,480 controls). For the asthma phenotype we identified individuals with the condition who had a positive response for self-reported code 20002_1111 (28,576 cases and 222,649 controls). For height we used 251,262 individuals in total with phenotype code 50.

To quantify the performance of the PRS for binary traits we calculated the area under the curve (AUC) (for binary traits) between the predicted and observed phenotypes using the R package “pROC”. Similarly, for quantitative traits we calculated the squared correlation between the PRS and the measured trait (r^2^). Table 1 summarises the cross-ancestry PRS evaluation parameters.

## Results

### Simulations of different trait, same-ancestry datasets

We performed simulations utilising common SNPs (MAF>1%) genotyped in the UK Biobank^26^ (UKBB) cohort. We compared shaPRS to two baseline approaches: single dataset analysis (*β*_1_ at all SNPs) and inverse variance weighted meta-analysis (*β*_12_ at all SNPs). The meta-analysis is equivalent to running shaPRS if there was no heterogeneity of effect anywhere across the genome, allowing us to quantify the extent to which incorporating the measure of heterogeneity (lFDR) learned via the Cochran test improves PRSs. In recent years, several methods that exploit genetic correlation between related traits to improve association or prediction accuracies have been proposed, including SMTPred^20^, MTAG^19^ and CTPR^53^. We choose SMTPred and MTAG as comparison methods because they also rely on genome-wide summary statistics and thus have a similar use-case to shaPRS. However, like other previously developed methods, both SMTPred and MTAG assume a constant shared genetic aetiology across the genome. A detailed description of the simulation can be found in the Materials and Methods section.

Genetic correlation (rG), which is a scalar metric, does not fully capture the overall structure of shared genetic aetiology. For example, a genetic correlation of 0.5 can be the result of all causal SNPs shared with a per-SNP effect correlation of 0.5 or, alternatively, only half of the causal SNPs may be shared but with an effect correlation of 1.0. By fixing the genetic correlation at 0.5, but varying the fraction of shared and non-shared genetic effects we investigated and demonstrated the key ability of our method to adapt to such different compositions of overlapping genetic aetiologies. We also considered an additional scenario, where five SNPs contribute 5% of the total non-shared heritability for each trait. The rationale for including such SNPs was to model highly penetrant variants such as *NOD2* in CD^25^ or *FLT3* in autoimmune thyroid disease^24,54^, which play an important role in differentiating these diseases from otherwise genetically related conditions. Our main simulation analyses examined 162 different genetic architectures that arose from the examined parameters. The full set of parameters are summarised in Table S1, and Fig 1 presents a subset of our simulation results with a heritability of 0.5, 1,000 causal variants and an rG of 0.5 between the proximal and adjunct datasets. The full set of results from all simulation scenarios can be found in Figs S3 and S4.

**Fig 1:**
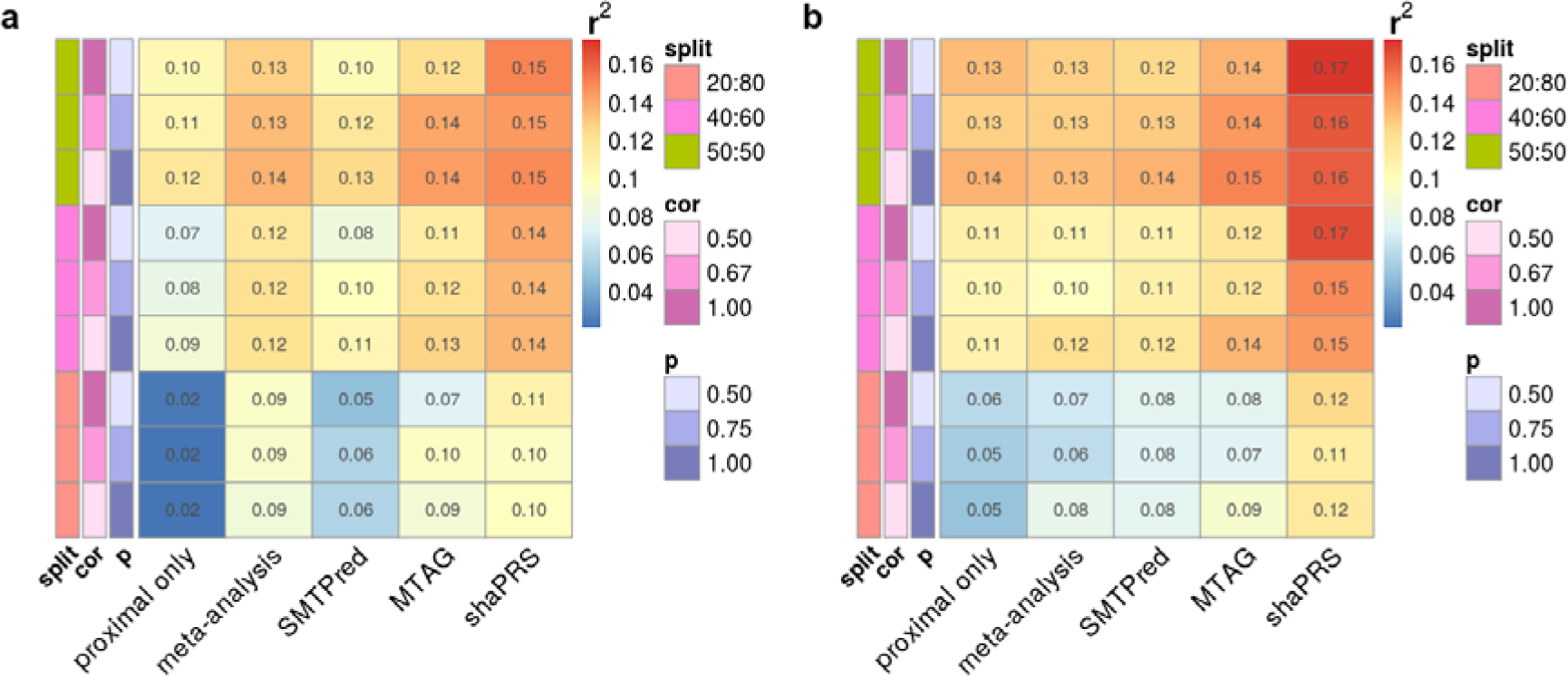
Heatmap of the squared correlation between simulated and predicted phenotypes for selected cross-trait genetic relationships. Warmer colours indicate better performance. **a.** A genome-wide genetic correlation between proximal and adjunct traits of 0.5 with a heritability of 0.5 from a 1,000 causal variants and no extra heterogeneity created by SNPs of large effect. Sample size N = 14,044, with a proximal/adjunct sample ratio of 50/50, 40/60 or 20/80, and where *cor* is the correlation of effect sizes between SNPs and *P (or causal_S_)* is the fraction of causal SNPs shared between the proximal and adjunct datasets,. *split* is the ratio of the proximal to adjunct dataset sizes. **b.** The same scenario as **a**, with the addition of extra heterogeneity created by five SNPs of large effect that contributed 5% non-shared heritability. Results across the complete set of simulated scenarios are shown in Fig S3.

shaPRS outperformed alternative methods in 93% of the simulated scenarios, and frequently by large margins. Fig S6 visualises the formal evaluation (via *r2redux’ r2_diff* function) between shaPRS and other methods as a heatmap for the scenarios depicted in Fig 1. ShaPRS’ capacity to accommodate genetic heterogeneity at a per-SNP level was particularly demonstrated by by an increasingly larger performance advantage over other methods in scenarios (Fig S1) where a given genetic correlation between two traits was concentrated amongst a subset of causal SNPs with stronger effect size correlations (See rG composition in Table S1). Reassuringly, shaPRS performed similarly to other methods in scenarios with a constant shared genetic aetiology (all causal SNPs shared between traits with weaker correlation in effect sizes). The relative ordering of the performance of the methods did not change with the introduction of extra heterogeneity created by SNPs of large effect (Fig 1b and Fig S1b). However, such high penetrance variants further enhanced the advantage of shaPRS against all evaluated alternatives. In conclusion, shaPRS compared favourably to all other approaches, particularly in scenarios when the underlying assumption of no non-shared SNPs with non-null effects was violated.

### Application to inflammatory bowel disease subtypes

Inflammatory bowel disease (IBD) is a complex inflammatory disease of the gastrointestinal tract with a prevalence of 0.5% in Western countries^55^. Its two main clinical subtypes, Crohn’s disease (CD) and ulcerative colitis (UC) have a substantial but imperfect overlap in their genetic aetiologies, with a genome-wide genetic correlation of ∼0.56^56^. We performed a shaPRS analysis of ulcerative colitis (UC) and Crohn’s disease (CD) using a GWAS dataset^35^ that included 4,647 and 5,400 UC and CD cases, respectively, and 10,308 shared controls. The Manhattan plot in Fig 2a illustrates how the estimated lFDR values capture the landscape of heterogeneity between UC and CD, with areas of highly incongruent effects (such as the *NOD2* region on chromosome 16) featuring prominently among the peaks.

**Fig 2:**
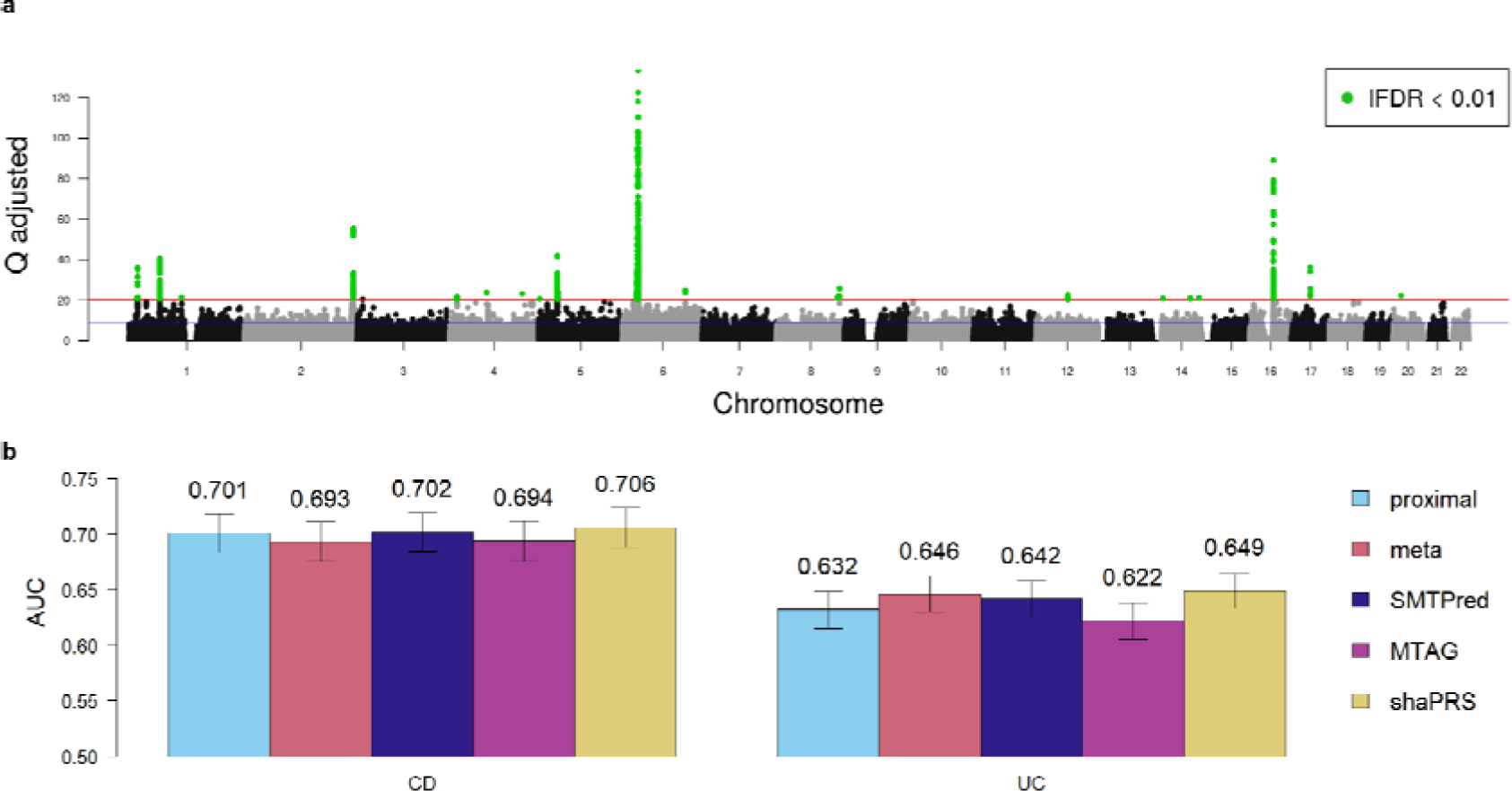
Comparison of PRS estimation methods for predicting inflammatory bowel disease subtypes. **a**. Manhattan plot depicting the genome-wide heterogeneity between Crohn’s disease and ulcerative colitis measured by Cochran’s Q test (Y-axis). Blue line represents SNPs with an lFDR < 0.5 and the red line represents SNPs with an lFDR < 0.01, which are also highlighted in green. **b.** Barplot of PRS performance evaluated by the area under the receiver operating characteristic curve (AUC) of the predicted and observed phenotypes in independent cohorts of 1,181/2,896 and 1,909/2,764 cases/controls, for Crohn’s disease (CD) and ulcerative colitis (UC), respectively. Each coloured bar represents a different PRS estimation method: IBD subtype alone (proximal: cyan), fixed-effect meta analysis (meta: orange), SMTPred (dark blue), MTAG (pink) and shaPRS (yellow). The error bars represent the 95% confidence intervals which were computed with 2,000 stratified bootstrap replicates and the values above each bar show the AUC for the given method.

A set of three baseline PRS were built, trained either on summary statistics from a case/control GWAS of a single disease subtype (CD or UC alone), or alternatively from a fixed-effect meta analysis of the CD and UC GWAS summary statistics to create a GWAS for the IBD phenotype). Three additional PRS were built based on more advanced models implemented in SMTPred, MTAG and shaPRS. All PRS were built using PRS-CS. We evaluated PRS performance on independent CD^26,37^ and UC^36^ cohorts, with 1,181/2,896 and 1,909/2,764 cases/controls, respectively, by estimating the squared correlation between the predicted and observed phenotypes (Fig 2 and Table S3).

We found that the PRS estimated from either the GWAS of CD or UC predicted the corresponding subtype with similar accuracy to the PRS generated from the IBD fixed-effect meta-analysis. Considering the variance-bias trade-off latent in these experiments, these results make intuitive sense; we approximately doubled the sample size of the cases for traits that share approximately half their genetic aetiology (rG=0.56). Therefore, given this level of shared genetic aetiology, combining phenotypes to train PRS did not consistently improve the accuracy. However, we found that shaPRS substantially outperformed these baseline PRS. Evaluated against the proximal dataset alone, shaPRS improved results by ∼4% and by ∼22%, for CD and UC, respectively. Compared to combining the CD and UC phenotypes in a fixed-effect meta analysis, shaPRS increased performance by ∼12% and by ∼6%, for CD and UC, respectively. Additionally, shaPRS also outperformed SMTPred by ∼7% and ∼10% and MTAG by ∼11% and 39%, for CD and UC, respectively. ShaPRS was significantly better than both SMTPred and MTAG, by at least either the *r2redux r2_var* or the Delong tests (Table S4). We also found that adding shaPRS into a nested model of the other method always improved the overall model fit, whereas the other way around, adding the other method into a nested shaPRS-only model, only improved the model fit once (in the case of MTAG for CD).

### Leveraging datasets from different ancestries

To date, GWAS have been predominantly focussed on European populations. The accuracy of PRS generated from GWAS summary statistics in one ancestry is decreased in individuals of other ancestries due to a combination of differences in LD, MAF, and causal variant effects between populations. We hypothesised that shaPRS could better leverage information from GWAS in different ancestries to construct more performant PRS. To test this hypothesis, we obtained GWAS summary statistics across a range of traits from diverse population pairs (Table 1, Materials and Methods), and quantified the extent to which shaPRS’ use of the adjunct data improved the performance of two PRS methods (PRS-CS and LDpred2) that only make use of GWAS summary statistics from the proximal ancestry. These methods require LD matrices representative of the study population, and are therefore restricted to single population analysis. We present their standard results as baselines, as well as the results with shaPRS pre-processing to leverage information from the adjunct datasets. Both methods can either use an additional validation dataset to optimise parameters, or estimate these internally using an “auto” mode. We used the auto mode to focus on the situation where an independent set of summary statistics is unavailable for the proximal population (which we believe will predominantly be the case for understudied populations). We also compared shaPRS against PRS-CSx^23^, a recently developed method that integrates GWAS summary statistics across different ancestries while accounting for MAF and LD differences. PRS-CSx is performed in two stages. Stage 1 infers posterior SNP effect sizes under continuous shrinkage (CS) priors, learnt either directly from the proximal dataset (*auto* mode) or optimised using a second independent set of GWAS summary statistics from the same proximal population. We again chose to use the *auto* approach, and refer to this as PRS-CSx-stage1. Note that this is not the recommended way to run PRS-CSx, and we include stage1 only to assess the performance without any additional dataset. Finally, to quantify the added value an independent proximal dataset brings to PRS performance, over and above any provided by shaPRS, we provided PRS-CSx with an independent dataset over which to optimise the weighted averaging of effects between the proximal and adjunct ancestries, which we refer to as PRS-CSx. The performance of each PRS method was evaluated by estimating the r^2^ and area under the curve (AUC) (for binary traits) between the predicted and observed phenotypes (Fig 3 and Table S2). We have also performed formal significance tests and likelihood-ratio tests that compared shaPRS and the other method baselines, relative to a complex model that had both (Table S5).

**Fig 3:**
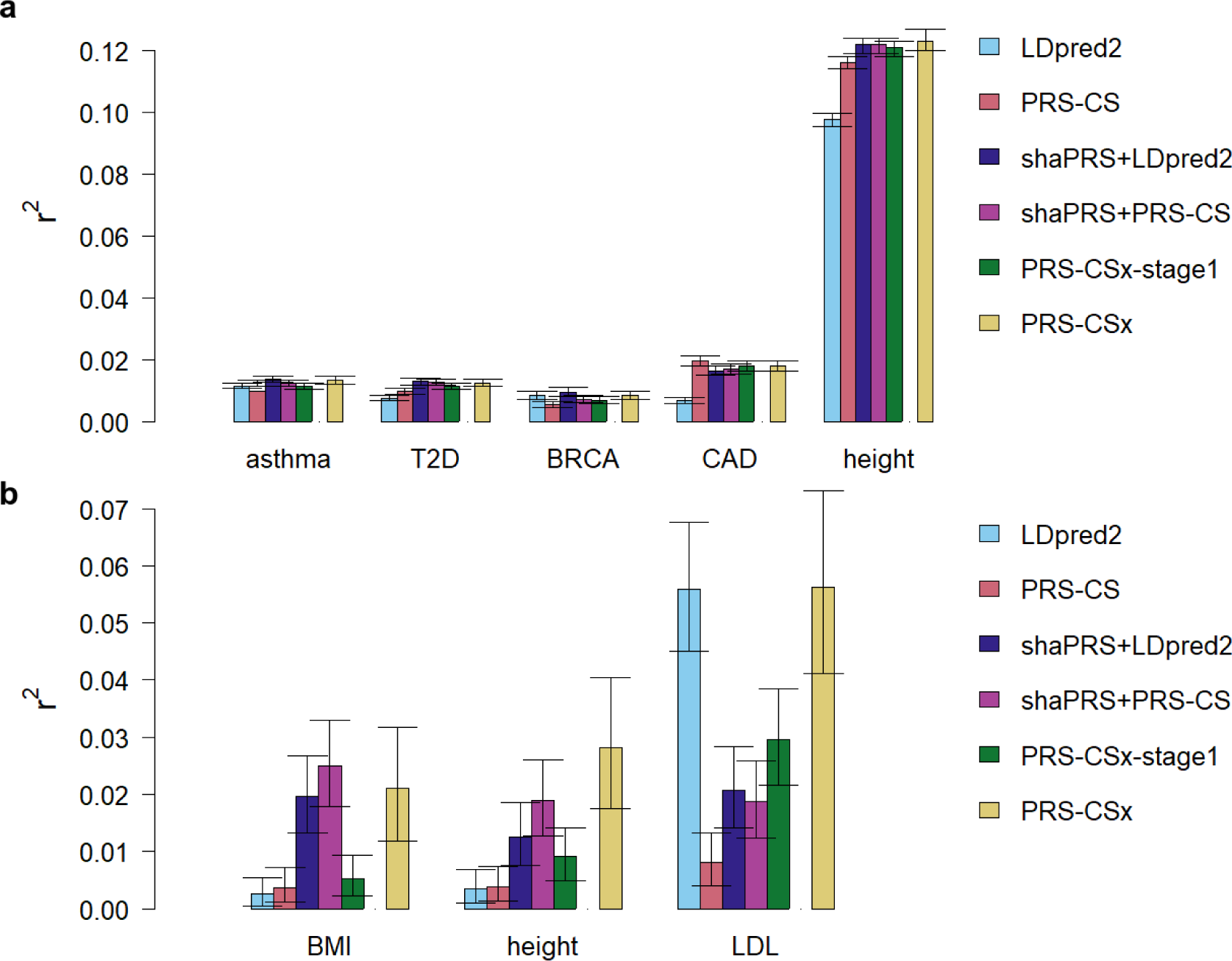
shaPRS maximises the accuracy of polygenic risk scores across divergent ancestry groups when only GWAS summary statistics are available. *LDpred2* and *PRS-CS* trained on only the proximal ancestry datasets. *shaPRS+LDpred2* and *shaPRS+PRS-CS* add preprocessing by shaPRS to leverage the adjunct datasets whilst generating a proximal ancestry PRS. *PRS-CSx-stage1* combines the proximal and adjunct summary data without the reliance on additional genotype validation data. *PRS-CSx* follows on from *PRS-CSx-stage1* by performing an additional step of finding the best linear combination of the proximal and adjunct PRS files by using additional genotype validation data. As *PRS-CSx* assumes a different use-case than our paper, it is only included as a point of reference. **a**. Barplot of PRS performance for the EUR proximal and EAS adjunct analyses, evaluated by the squared Pearson correlation coefficient (**r^2^**) between predicted and observed phenotypes. Confidence intervals were generated via the *r2redux’ r2_var* function. T2D = type 2 diabetes, BRCA = breast cancer, CAD = coronary artery disease. **b**. Barplot of PRS performance for the AFR proximal and EUR adjunct analyses, evaluated by the squared Pearson correlation coefficient (**r^2^**) between predicted and observed phenotypes. BMI = body mass index, LDL = low-density lipoprotein cholesterol level.

ShaPRS improved the accuracy of PRS estimates from LDpred2 and PRS-CS in six of the eight traits studied, with the greatest improvement seen for traits where the power of the adjunct dataset far outweighed that of the proximal study (e.g. BMI and height in the African_proximal_ and European_adjunct_ studies). shaPRS also consistently outperformed PRS-CSx-stage1 in these same six studies, in four instances also reaching statistical significance (Table S4), demonstrating its superiority in situations where only a single set of GWAS summary statistics are available for a given proximal population. Furthermore, the shaPRS improved PRSs were more performant than those from PRS-CSx for four of the eight traits studied, despite the fact that PRS-CSx exclusively made use of an independent dataset from the proximal population. PRS-CSx only outperformed shaPRS improved scores by a noticeable margin for two of the eight tested traits (height and LDL in the African_proximal_ and European_adjunct_ studies), highlighting the extent to which shaPRS can improve PRS without the need for a second independent set of GWAS summary statistics from the proximal population.

To better understand how shaPRS gains performance, we examined two analyses in detail. Taking the Asthma analysis in EUR/EAS individuals, and the CD/UC analysis as examples, we can see that very few SNPs are detected to have genuinely different effects (i.e. low lFDR) in the different ancestries or traits (Figure 4a-b). Amongst SNPs with low heterogeneity (Figure 4c), shaPRS tends to shrink the effect size, beta, towards zero for SNPs ultimately declared non-significant, while leaving beta on average unchanged for significant SNPs. For all low heterogeneity SNPs, the standard error shrinks as would be expected in any meta analysis. For high heterogeneity SNPs (Figure 4d), on the other hand, both beta and the standard error remain relatively unchanged regardless of significance. shaPRS thus allows effects specific to individual traits or populations to be leveraged when appropriate, and does not attempt to leverage that information when inappropriate. Given the predominance of low heterogeneity, non-significant SNPs, the greatest effect of shaPRS is to shrink estimates of null SNPs towards zero.

**Fig 4.**
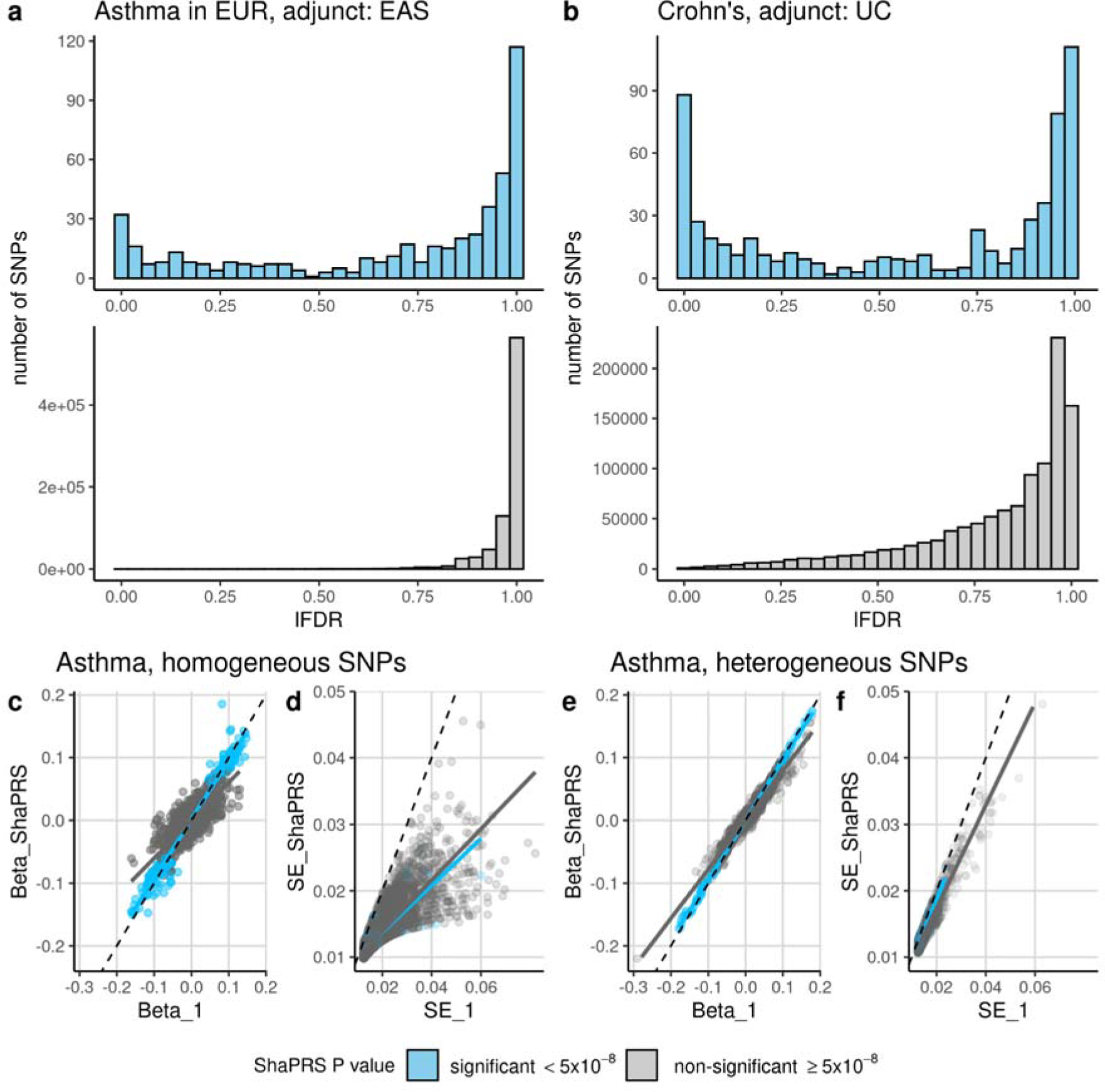
shaPRS maximises accuracy of polygenic risk scores by more completely capturing between SNP variation in shared effects across ancestry groups or related traits. . The top row contrasts the distribution of effect heterogeneity measured by lFDR in **a)** a cross-ancestry analysis of asthma that uses European GWAS summary statistics as the proximal dataset and East Asian GWAS summary statistics as the adjunct dataset (left), and **b)** a cross-trait analysis of Crohn’s disease, leveraging a GWAS of ulcerative colitis (UC) as an adjunct dataset. The distributions of lFDR values are shown, where low lFDR corresponds to higher heterogeneity in estimated effects. The bottom row compares the proximal study’s beta (Beta_1) and standard error (SE_1) to its shaPRS-adjusted output (Beta_shaPRS, SE_shaPRS respectively) for the asthma analysis. SNP have been partitioned into those with little evidence of heterogeneity (lFDR > 0.5) between the two populations (**c**, **d**) and this with evidence of heterogeneity (lFDR <= 0.5) between the two populations(**e**, **f**). Colours indicate whether a SNP was detected to have a significantly non-zero effect (p < 5x10^-8^) in the shaPRS analysis.

## Discussion

We introduce shaPRS, a novel method that integrates genetic association information from heterogeneous sources to improve accuracy of PRS for related traits and across ancestral populations.

A major strength of shaPRS is the ability to exploit the differential genetic architecture of related traits by considering the evidence for heterogeneity at each variant and weighting towards the estimate with the more beneficial properties: smaller variance in case of low heterogeneity or, alternatively, smaller bias in case of high heterogeneity. shaPRS can thus particularly improve the accuracy of a PRS when the genetic correlation structure between the proximal and adjunct datasets varies between SNPs. This is in contrast with previously developed methods, such as SMTPred and MTAG, both of which assume a constant sharing of genetic aetiology. SMTPred assumes that all SNP effects are shared equally (it integrates predictions on the PRS-level), whereas MTAG learns different per-SNP weights, however, unlike shaPRS, the combined SNP effects are a function of parameters that are constant across all variants. The per-SNP weighting approach adopted by shaPRS requires a given SNP to be genotyped in both the proximal and adjunct studies, otherwise the effect from the proximal study alone is used to generate the PRS. The advantage of applying shaPRS pre-processing is therefore positively correlated with the number of SNPs genotyped in both the proximal and adjunct datasets. Given the widespread use of a small number of genotyping arrays and genotype imputation, we expect most traits or ancestries to have sufficient overlapping SNPs to make shaPRS pre-processing a useful strategy.

In our example of Crohn’s disease and ulcerative colitis, the pervasive sharing of genetic effects between the two diseases is well established^57^, and the genetic correlation between the two diseases has been estimated to be 0.56^56^. However, there are some SNPs with large differences in effect between Crohn’s and UC^57^; for example, in the *NOD2* locus genetic variants explain around 1.5% of variance in liability of Crohn’s disease^58^, but there is no evidence of association to ulcerative colitis. More fully accounting for this inconsistent correlation in genetic effects between traits enables shaPRS to outperform competing cross-trait methods. In the case of Crohn’s disease risk prediction, shaPRS outperformed fixed-effect meta analysis, SMTPred and MTAG by ∼12%, ∼7% and ∼11%, respectively (see Table S3 for detailed results).

When applying our method to cross-ancestry prediction based solely on GWAS summary statistics, shaPRS with either LDpred2 or PRS-CS, outperformed the cross-ancestry method PRS-CSx for six of the eight traits considered. Even when we exclusively provided PRS-CSx with this additional genotyped dataset to fine tune the final PRS by finding the best linear combination of the proximal and adjunct PRS, it only appreciably outperformed a shaPRS-informed analysis for two of the eight tested traits. We note that such a fine-tuning approach could, in principle, be applied to the outputs of any PRS method (including those built by shaPRS) to further improve their PRS accuracy across populations. Unfortunately, we believe that additional genotyped datasets will seldom be available for most underrepresented populations and lower prevalence traits, making shaPRS ability to generate accurate PRS without these a key feature of our method.

Comparing the full with the nested models of either just shaPRS or just one of the other methods, also demonstrated our method’s advantages, as evidenced by the consistently lower likelihood-ratio test p-values favouring shaPRS. This held true for both the simulations (Fig S7) as well as for the majority of the real data analyses (Tables S4 and S5), despite the limited sample sizes and therefore lower power for the latter. We interpret these results to suggest that shaPRS was able to achieve a higher performance by adding unique information not available to the other methods.

In the coming years, to expand the clinical applicability of PRS, more ancestrally diverse populations will need to be recruited for large-scale genetic research^12,13^. In the interim, methods such as shaPRS can contribute to more equitable health outcomes by leveraging existing datasets more effectively. Our simulations and real-world examples show that shaPRS can improve PRS estimation across a broad range of genetic architectures. While we have showcased the power of shaPRS for improving PRS estimates between traits and ancestries, this flexibility enables shaPRS to be applied whenever incomplete sharing of genetic effects is expected between two GWAS datasets. Other possible use cases for shaPRS could therefore include generating PRS for traits with heterogeneity of effect between the sexes or between different environments.

ShaPRS is designed to fit within existing pipelines as a pre-processing tool, thus, it is not in direct competition with other PRS generation tools such as LDpred2^29^ or PRS-CS^30^. shaPRS can therefore continue to be applied as more performant PRS methods are derived in the future, such as the recently proposed PolyPred^59^.Our recommended approach is to pre-process GWAS summary statistics via shaPRS before taking them forward to a PRS tool of choice that would be used to produce the final profile scores. shaPRS also fits with the ongoing trend of reliance on summary statistics alone, without the need for access to genotype level data at any stage, as it provides a competitive performance without the need for a validation genotype cohort. Our method is open source and is freely available from https://github.com/mkelcb/shaprs.

## Declaration of interests

C.A.A. has received consultancy or lectureship fees from Genomics plc,BridgeBio Inc and GSK. C.W. receives funding from GSK and MSD and is a part time employee of GSK. These companies had no input into this work

## Funding information

This work was funded by the Wellcome Trust (220540/Z/20/A, ‘Wellcome Sanger Institute Quinquennial Review 2021-2026’, 203950/Z/16/A, WT220788, WT107881, 206194, 108413/A/15/D) and the MRC (MC_UU_00002/4) and supported by the NIHR Cambridge BRC (BRC-1215-20014). The views expressed are those of the author(s) and not necessarily those of the NHS, the NIHR or the Department of Health and Social Care. For the purpose of Open Access, the author has applied a CC BY public copyright licence to any Author Accepted Manuscript version arising from this submission.

This research was conducted using the UK Biobank Resource under Application Number 30931.

## Data and code availability

shaPRS R package is available from https://github.com/mkelcb/shaprs. Code to perform all analyses reported in this manuscript is available at https://github.com/mkelcb/shaprs-paper. The final PRS files and diagnostic data are available from the Supplementary data. The Crohn’s disease and ulcerative colitis genotype data used here can be obtained via managed access at: https://ega-archive.org/studies/EGAS00001000924, https://ega-archive.org/studies/EGAS00000000084 and https://ega-archive.org/datasets/EGAD00000000005. The standard LD reference panels for LDpred2 and PRS-CS may be obtained from https://figshare.com/articles/dataset/European_LD_reference_with_blocks_/19213299 and https://personal.broadinstitute.org/hhuang//public//PRS-CSx/Reference/, respectively.

## Acknowledgements

We thank all individuals who donated or collected samples used in this study.

## Notes

### Competing Interest Statement

C.A.A. has received consultancy fees from Genomics plc and BridgeBio Inc. C.W. receives funding from GSK and MSD.

### Funding Statement

This work was funded by the Wellcome Trust (203950/Z/16/A, WT220788, WT107881, 206194, 108413/A/15/D) and the MRC (MC_UU_00002/4) and supported by the NIHR Cambridge BRC (BRC-1215-20014). The views expressed are those of the author(s) and not necessarily those of the NHS, the NIHR or the Department of Health and Social Care. For the purpose of Open Access, the author has applied a CC BY public copyright licence to any Author Accepted Manuscript version arising from this submission.
This research was conducted using the UK Biobank Resource under Application Number 30931.

### Author Declarations

ShaPRS R package is available from https://github.com/mkelcb/shaprs. Code to perform all analyses reported in this manuscript is available at https://github.com/mkelcb/shaprs-paper. The final PRS files and diagnostic data are available from the Supplementary data. The Crohn's disease and ulcerative colitis genotype data used here can be obtained via managed access at: https://ega-archive.org/studies/EGAS00001000924, https://ega-archive.org/studies/EGAS00000000084 and https://ega-archive.org/datasets/EGAD00000000005.

### Summary of Updates

We have enacted the most important suggestions of the reviewers, which have further demonstrated the power of shaPRS for building highly performant polygenic risk scores across traits and ancestries. These changes include, but are not limited to: 1.Formally evaluating the difference in performance between shaPRS and other methods 2.Demonstrating that adding shaPRS into a baseline model contributes non-overlapping information. 3.Adding confidence intervals for the continuous trait PRS.

