## Supplementary material for "ShaPRS: Leveraging shared genetic effects across traits or ancestries improves accuracy of polygenic scores"

*These authors contributed equally

**shaPRS walkthrough examples**

The following three examples may help to explain the application of shaPRS in three illustrative scenarios of SNPs: fully shared, non-shared and partially shared effect between studies.

**Fully shared SNP.** A SNP whose effect is 100% shared between proximal and adjunct studies. These are frequently null SNPs that have a true effect size of 0 ($\beta_{1}=\beta_{2}=\beta_{12}=0$). Here, the estimated lFDR would be close to 1 ($\pi$=1), so the shaPRS equation

$\beta_{shaPRS}= \left( 1-\pi\right)\beta_{1} + \pi\beta_{12}$,

would simplify to

$\beta_{shaPRS}⋍ 1 * \beta_{12}$.

Thus here the final SNP estimate would become close to identical to the meta-analysis ($\beta_{12}$).

**SNP specific to proximal study.** A SNP that only has an effect in the proximal study, but not in the adjunct study. An example of this would be some SNPs in the *NOD2* region in our IBD analyses, which has an effect only on CD susceptibility. Here, the estimated lFDR would be 0 ($\pi$=0). Therefore, the shaPRS equation would simplify to

$\beta_{shaPRS}⋍ 1 * \beta_{1}$.

Thus here the final SNP estimate would become close to the proximal study ($\beta_{1}$).

**SNP partially shared between studies.** A SNP that has an effect on both phenotypes, but with different effects in proximal and adjunct studies (which should give rise to a low lFDR for the variant’s Cochran’s test). Here, the estimated lFDR would be x ($\pi$=x), which would be a value between zero and one. Therefore, the shaPRS equation would become

$\beta_{shaPRS}= \left( 1-x \right)\beta_{1} + x \beta_{12}$.

Which would make $\beta_{shaPRS}$ take on an intermediate value between the proximal and the meta-analysis that depends on the exact degree of effect sharing, specific to each SNP.

**Missing SNPs and practical application of shaPRS**

By default, the shaPRS R package will keep SNPs that are missing in the adjunct data by using their proximal data estimates, which is expected to produce the best overall quality PRS in practical applications. However, for our comparisons we excluded all SNPs that were missing from the adjunct datasets to ensure all methods worked off the same set of SNPs. Incorporating estimates from proximal-only SNPs would have had the effect of adding a constant value to all methods' PRS, which would not have altered the rank order of the methods.

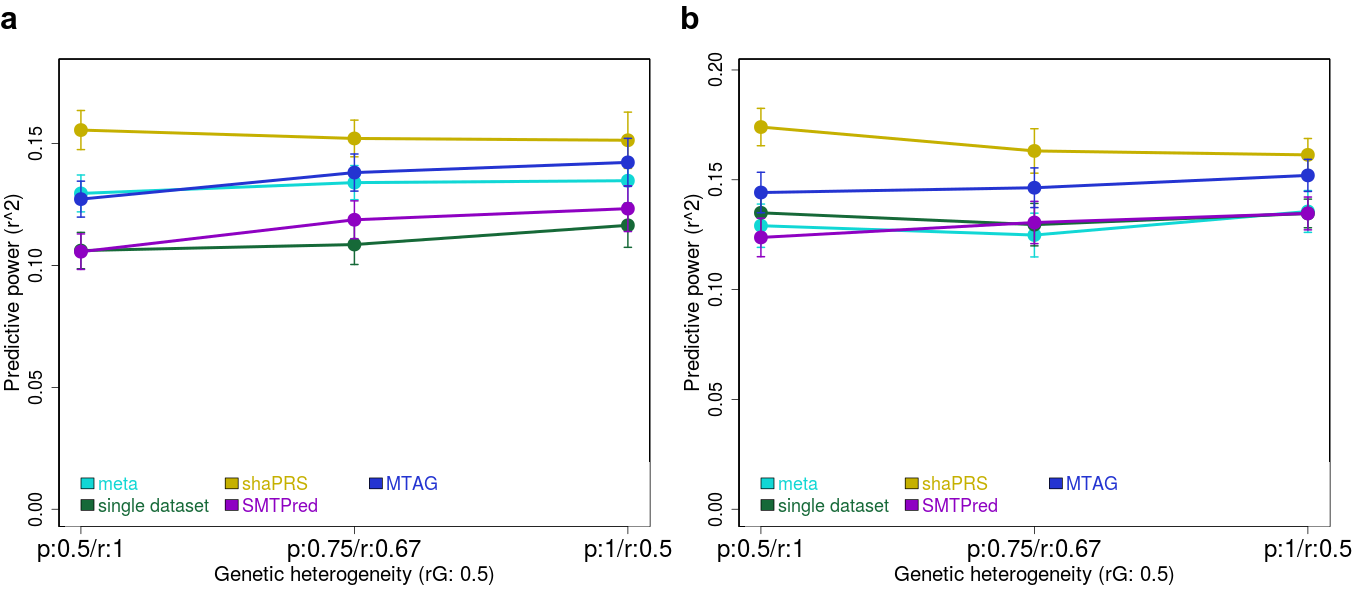

**S Fig 1 a.** The effect of varying the composition of heterogeneity in the genetic correlation (the proportion of shared causal effects to their correlation) across the five methods. The X-axis shows the three different compositions that were used to generate the same genetic correlation (rG = 0.5). The axis labels are coded as *p/r*, which are the shared fraction of causal SNPs / effect size correlation of these SNPs. Y-axis is the squared correlation between the predicted and observed phenotypes on the test set and the error bars represent the standard error of mean. Meta-analysis (blue) represents the PRS built from combining both phenotypes. Single dataset (green) represents PRS built from only the individuals from the proximal dataset. shaPRS (yellow) is our method, MTAG (blue) is a baseline method that generates PRS via estimate SNP effect sizes based on constant parameters, and SMTPred (purple) is the baseline method that produced a PRS by balancing the PRS for proximal and adjunct datasets based on their genetic correlation. **b.** The same scenario as **a**, with the addition of the extra heterogeneity created by SNPs of large effect that contributed 5% non-shared heritability.

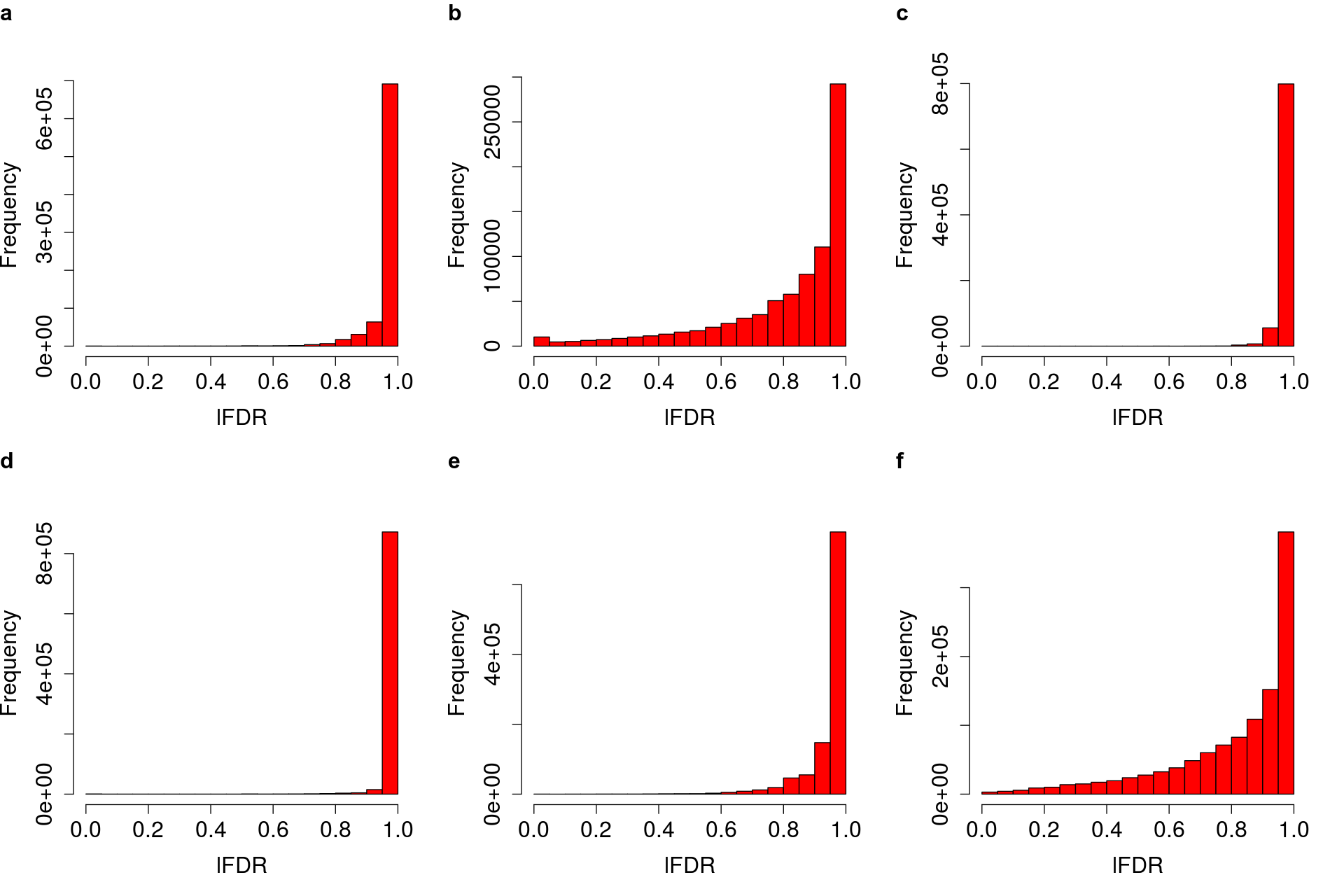

**S Fig 2: a, b, c, d**, **e** and **f** Histograms of lFDR of heterogeneity between ancestries for asthma, height, BRCA, CAD, and T2D and between UC/Crohn’s disease for IBD, respectively.

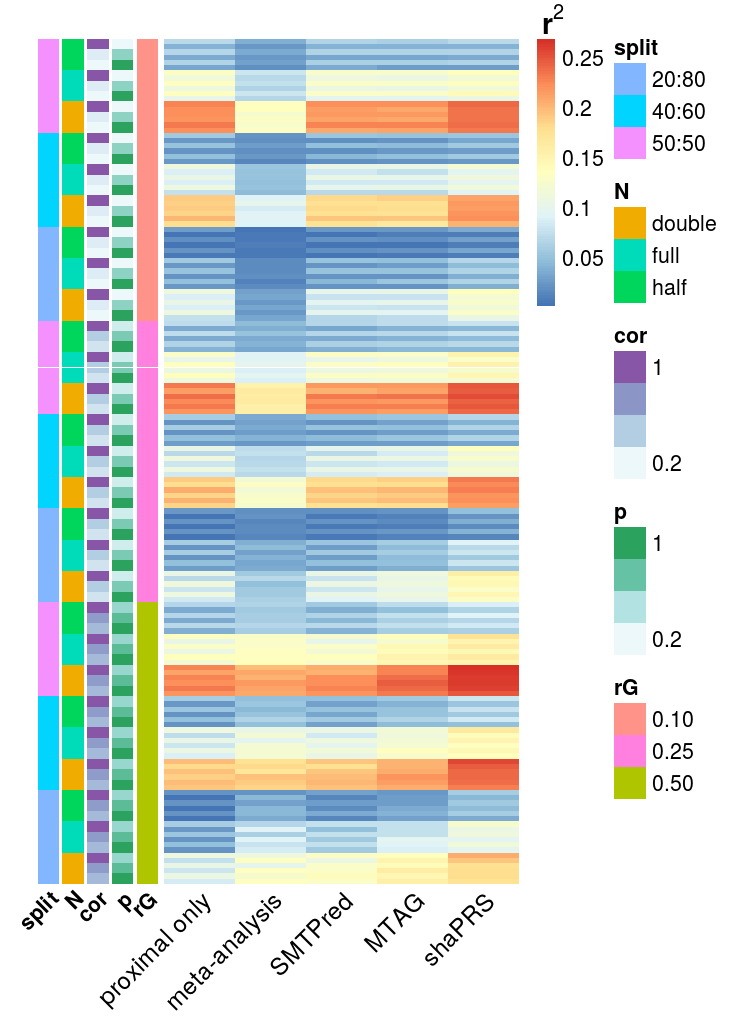

**S Fig 3:** Heatmap of results for all 162 simulation scenarios, colours indicate squared correlation between simulated and predicted phenotypes for example scenarios, where warmer values indicate better performance. *N* is the sample size, half, full and double are 7,022, 14,044 and 28,088 individuals, respectively. *p* is the fraction of causal SNPs shared between the proximal and adjunct datasets, *cor* is the correlation of effect sizes between these SNPs. *split* is the ratio of the proximal to adjunct dataset sizes. *rG* is the genetic correlation between the proximal to adjunct datasets.

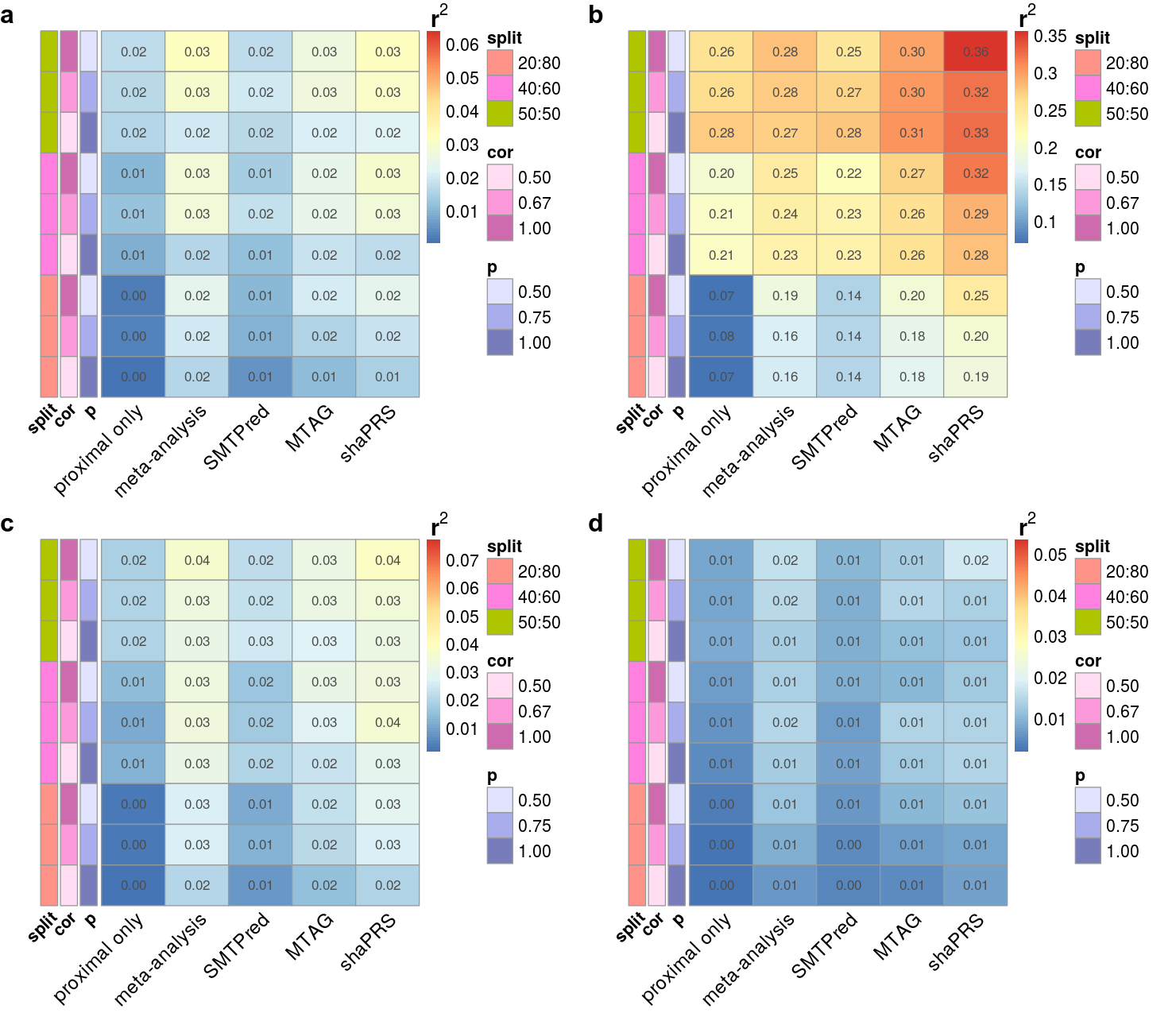

**S Fig 4:** Heatmap of the squared correlation between simulated and predicted phenotypes in the additional 36 scenarios exploring selected parameters. *p* is the fraction of causal SNPs shared between the proximal and adjunct datasets, *cor* is the correlation of effect sizes between these SNPs. *split* is the ratio of the proximal to adjunct dataset sizes. Warmer colours indicate better performance. **a.** Sample size N = 14,044, with a proximal/adjunct sample ratio of 50/50, 40/60 or 20/80, a genetic correlation between proximal and adjunct traits of 0.5 with a heritability of 0.25 from a 1,000 causal variants, no extra heterogeneity created by SNPs of large effect. **b.** The same scenario as **a**, with a heritability of 0.75. **b.** **c.** The same scenario as **b**, with a heritability of 0.5 and 3,000 causal SNPs. **d.** The same scenario as **c**, with a 5,000 causal SNPs.

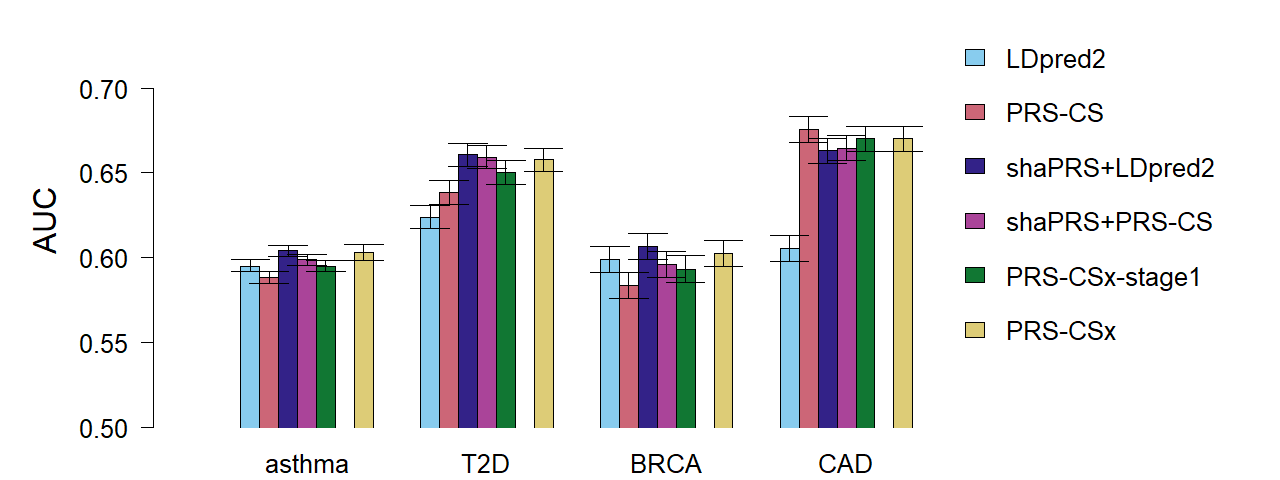

**S Fig 5:** **shaPRS maximises accuracy of polygenic risk scores across divergent ancestry groups when only GWAS summary statistics are available.** Barplot of the results of the cross-ancestry analysis that compared the accuracy of six different methods to produce a PRS. *LDpred2* and *PRS-CS* are the LDpred2 method on auto option and the PRS-CS method, both trained on only the target ancestry datasets. *shaPRS+LDpred2* and *shaPRS+PRS-CS* add preprocessing by shaPRS to leverage the adjunct datasets whilst generating a target ancestry-specific PRS. *PRS-CSx-stage1* combines the proximal and adjunct summary data without the reliance on additional genotype validation data. *PRS-CSx* follows on from *PRS-CSx-stage1* by performing an additional step of finding the best linear combination of the proximal and adjunct PRS files by using additional genotype validation data. As *PRS-CSx* assumes a different use-case than our paper, it is only included as a point of reference. Barplot for the EUR proximal with EAS adjunct scenario of PRS performance evaluated by the area under the receiver operating characteristic curve (AUC) of the predicted and observed phenotypes. The error bars represent the 95% confidence intervals which were computed with 2,000 stratified bootstrap replicates.

| **Table S1 \| Range of parameters evaluated in the simulation experiments.** | |
| --- | --- |
| **parameter** | **range** |
| sample size | 7,022, 14,044 and 28,088 training individuals |
| phenotype split (proximal/adjunct) | 50/50, 40/60 and 20/80 |
| five large effect SNPs | enabled or disabled |
| rG composition | **rG** \| **shared fraction of causal SNPs** \| **effect size correlation**; \| 0.1 \| 1; 0.1 \| 0.55 \| 0.182; \| 1 \| 0.1; \| 0.25 \| 1; 0.25 \| 0.625 \| 0.4; \| 1 \| 0.25; \| 0.5 \| 1; 0.5 \| 0.75 \| 0.667; \| 1 \| 0.5 |
| Sample size represents the number of individuals used for training the PRS, which were chosen to be approximately half , equal to or double the size of our UC GWAS datasets (N = 4,647 cases and 10,308 controls). Phenotype split represents the percentage of the samples with quantitative phenotypes simulated for each of the two traits, given as proximal/adjunct. The *‘five large effect SNPs’* represents the choice to include five highly penetrant SNPs that explained 5% of the non-shared heritability of each trait. *rG* composition represents the different ways genetic correlations were constructed as a product of three different ‘shared fraction of causal SNPs’ and ‘effect size correlation’ estimates. | |

| **Table S2 \| Leveraging information from GWAS studies with different ancestries** | | | | | | |
| --- | --- | --- | --- | --- | --- | --- |
| **Target ancestry** | **trait** | **data** | **information pooling** | **PRS** | **r^2^** | **AUC** |
| EUR | T2D | proximal (EUR) | *N/A* | LDpred2 | 0.0075 | 0.624 |
|  |  |  |  | PRS-CS | 0.00958 | 0.639 |
|  |  | proximal  + adjunct | PRS-CSx | PRS-CSx | 0.0125 | 0.658 |
|  |  |  |  | PRS-CSx-stage1 | 0.0114 | 0.650 |
|  |  |  | shaPRS | LDpred2 | **0.0129** | **0.661** |
|  |  |  |  | PRS-CS | 0.0127 | 0.659 |
| EUR | height | proximal | *N/A* | LDpred2 | 0.0976 | *N/A* |
|  |  |  |  | PRS-CS | 0.116 |  |
|  |  | proximal  + adjunct | PRS-CSx | PRS-CSx | **0.123** |  |
|  |  |  |  | PRS-CSx-stage1 | 0.121 |  |
|  |  |  | shaPRS | LDpred2 | 0.122 |  |
|  |  |  |  | PRS-CS | 0.122 |  |
| EUR | BRCA | proximal | *N/A* | LDpred2 | 0.00828 | 0.599 |
|  |  |  |  | PRS-CS | 0.00529 | 0.584 |
|  |  | proximal  + adjunct | PRS-CSx | PRS-CSx | 0.00836 | 0.602 |
|  |  |  |  | PRS-CSx-stage1 | 0.00684 | 0.593 |
|  |  |  | shaPRS | LDpred2 | **0.00955** | **0.607** |
|  |  |  |  | PRS-CS | 0.00724 | 0.596 |
| EUR | CAD | proximal | *N/A* | LDpred2 | 0.00666 | 0.605 |
|  |  |  |  | PRS-CS | **0.0195** | **0.676** |
|  |  | proximal  + adjunct | PRS-CSx | PRS-CSx | 0.0179 | 0.670 |
|  |  |  |  | PRS-CSx-stage1 | 0.0179 | 0.670 |
|  |  |  | shaPRS | LDpred2 | 0.0164 | 0.663 |
|  |  |  |  | PRS-CS | 0.0169 | 0.665 |
| EUR | asthma | proximal | *N/A* | LDpred2 | 0.0115 | 0.595 |
|  |  |  |  | PRS-CS | 0.0099 | 0.588 |
|  |  | proximal  + adjunct | PRS-CSx | PRS-CSx | 0.0134 | 0.603 |
|  |  |  |  | PRS-CSx-stage1 | 0.0113 | 0.595 |
|  |  |  | shaPRS | LDpred2 | **0.0136** | **0.604** |
|  |  |  |  | PRS-CS | 0.0123 | 0.599 |
| **Target ancestry** | **trait** | **data** | **information pooling** | **PRS** | **r^2^** | **AUC** |
| AFR | BMI | proximal | *N/A* | LDpred2 | 0.0025 | *N/A* |
|  |  |  |  | PRS-CS | 0.0037 |  |
|  |  | proximal  + adjunct | PRS-CSx | PRS-CSx | 0.0210 |  |
|  |  |  |  | PRS-CSx-stage1 | 0.0053 |  |
|  |  |  | shaPRS | LDpred2 | 0.0196 |  |
|  |  |  |  | PRS-CS | **0.0249** |  |
| AFR | height | proximal | *N/A* | LDpred2 | 0.0035 | *N/A* |
|  |  |  |  | PRS-CS | 0.0039 |  |
|  |  | proximal  + adjunct | PRS-CSx | PRS-CSx | **0.0282** |  |
|  |  |  |  | PRS-CSx-stage1 | 0.0091 |  |
|  |  |  | shaPRS | LDpred2 | 0.0126 |  |
|  |  |  |  | PRS-CS | 0.0039 |  |
| AFR | LDL | proximal | *N/A* | LDpred2 | 0.0559 | *N/A* |
|  |  |  |  | PRS-CS | 0.0082 |  |
|  |  | proximal  + adjunct | PRS-CSx | PRS-CSx | **0.0563** |  |
|  |  |  |  | PRS-CSx-stage1 | 0.0296 |  |
|  |  |  | shaPRS | LDpred2 | 0.0208 |  |
|  |  |  |  | PRS-CS | 0.0187 |  |

Table of the results of the cross-ancestry analysis that compared the accuracy of six different methods to produce a PRS. **Target ancestry** is the genetic ancestry of the target individuals on whom the final PRS was evaluated. **trait** is the phenotype evaluated. **data** is the summary statistic datasets used for training. Proximal is the GWAS conducted in the target ancestry and proximal+adjunct is the target and adjunct GWAS together. The adjunct GWAS was sourced from Japanese individuals in case of European target PRS, and European individuals in the case of African target PRS. **information pooling** is the method that was used to pool the information from the proximal and adjunct datasets. *N/A* is when no information pooling took place, *PRS-CSx* is the PRS-CSx method and *shaPRS* is the method presented in this paper. **PRS** is the method that was used to generate the final PRS profiles. *LDpred2* is the PRS generated by the LDpred2-auto method that used no additional genotype data. *PRS-CS* is the PRS generated by the PRS-CS method that used no additional genotype data. *PRS-CSx* is the PRS generated by the PRS-CSx method that used validation data to weigh between the target and adjunct PRS. *PRS-CSx-stage1* is the PRS generated by the PRS-CSx method that did not use validation data to weigh between the target and adjunct PRS. **r^2^** is the squared Pearson correlation coefficient between predicted and observed phenotypes. **AUC** is the area under the receiver operating characteristic curve of the predicted and observed phenotypes. All PRS were evaluated on strictly non-overlapping subsets of the UK Biobank.

| **Table S3 \| shaPRS performance relative to other methods in IBD subtypes** | | |
| --- | --- | --- |
| **Other method** | **CD** | **UC** |
| proximal | 0.103 (-4%) | 0.052 (-22%) |
| meta-analysis | 0.095 (-12%) | 0.061 (-6%) |
| SMTPred | 0.100 (-7%) | 0.059 (-10%) |
| MTAG | 0.096 (-11%) | 0.044 (-39%) |
| shaPRS | 0.107 *(N/A)* | 0.065 *(N/A)* |

Table of the results for the inflammatory bowel disease subtype analysis that shows the performance improvements achieved by shaPRS relative to other methods. The values in each row are r^2^, the squared Pearson correlation coefficient between predicted and observed phenotypes, followed by the percentage difference relative to shaPRS. **CD** is Crohn’s disease and **UC** is ulcerative colitis.

| **Table S4A \| CD - shaPRS comparison against other methods** | | | | |
| --- | --- | --- | --- | --- |
|  | **model difference** | | **LRT of nested vs complex model**  **(p-value)** | |
| **method** | **r2redux p** | **Delong p** | **other** | **shaPRS** |
| **SMTPred** | 0.041 | 0.23 | 0.055 | 2.290E-09 |
| **MTAG** | 0.053 | 0.041 | 4.550E-07 | 3.69E-18 |

| **Table S4B \| UC- shaPRS comparison against other methods** | | | | |
| --- | --- | --- | --- | --- |
|  | **model difference** | | **LRT of nested vs complex model**  **(p-value)** | |
| **method** | **r2redux p** | **Delong p** | **other** | **shaPRS** |
| **SMTPred** | 0.029 | 0.044 | 0.171 | 1.530E-08 |
| **MTAG** | 2.440E-06 | 5.310E-06 | 0.138 | 1.200E-24 |

Results from the formal evaluation of model difference between shaPRS and other methods for the inflammatory bowel disease subtype. The **model difference** column shows the p-values if there was a difference between shaPRS and the other methods via the ‘r2redux r_diff’ and the pROC’ Delong’ tests, respectively. The **LRT of nested vs complex model** column shows the p-values for a likelihood ratio tests that evaluate if adding **shaPRS** or the **other** (non-shaPRS) PRS onto a nested model improves over the complex model of shaPRS+other. **A.** Crohn's disease and **B.** is Ulcerative Colitis.

| **Table S5A \| EUR-EAS asthma - shaPRS comparison against other methods** | | | | |
| --- | --- | --- | --- | --- |
|  |  | **method** | **PRS-CSx** | **PRS-CSx-stage1** |
| **model difference** | **shaPRS-PRSCS** | **r2redux p** | 9.420E-07 | 0.0026 |
|  |  | **delong p** | 3.290E-06 | 0.00257 |
|  | **shaPRS+LDpred2** | **r2redux p** | 0.047 | 4.83E-13 |
|  |  | **delong p** | 0.0947 | 5.64E-12 |
| **LRT of nested vs complex model** | **shaPRS-PRSCS** | **other** | 2.11E-37 | 7.420E-09 |
|  |  | **shaPRS** | 0.00296 | 7.12E-32 |
|  | **shaPRS+LDpred2** | **other** | 2.02E-16 | 3.260E-07 |
|  |  | **shaPRS** | 9.32E-34 | 3.28E-81 |

| **Table S5B \| EUR-EAS height - shaPRS comparison against other methods** | | | | |
| --- | --- | --- | --- | --- |
|  |  | **method** | **PRS-CSx** | **PRS-CSx-stage1** |
| **model difference** | **shaPRS-PRSCS** | **r2redux p** | 1.100E-03 | 0.387 |
|  | **shaPRS+LDpred2** | **r2redux p** | 0.821 | 0.127 |
| **LRT of nested vs complex model** | **shaPRS-PRSCS** | **other** | 2.66E-77 | 1.870E-56 |
|  |  | **shaPRS** | 7.05E-34 | 6.50E-69 |
|  | **shaPRS+LDpred2** | **other** | <2.225074e-308 | 2.600E-297 |
|  |  | **shaPRS** | <2.225074e-308 | <2.225074e-308 |

| **Table S5C \| EUR-EAS T2D - shaPRS comparison against other methods** | | | | |
| --- | --- | --- | --- | --- |
|  |  | **method** | **PRS-CSx** | **PRS-CSx-stage1** |
| **model difference** | **shaPRS-PRSCS** | **r2redux p** | 0.533 | 6.567e-06 |
|  | **shaPRS+LDpred2** | **r2redux p** | 0.219 | 2.359e-06 |
| **LRT of nested vs complex model** | **shaPRS-PRSCS** | **other** | 1.114e-17 | 1.106e-11 |
|  |  | **shaPRS** | 1.588e-24 | 6.481e-56 |
|  | **shaPRS+LDpred2** | **other** | 1.305e-28 | 1.729e-16 |
|  |  | **shaPRS** | 2.942e-43 | 1.816e-68 |

| **Table S5D \| EUR-EAS CAD- shaPRS comparison against other methods** | | | | |
| --- | --- | --- | --- | --- |
|  |  | **method** | **PRS-CSx** | **PRS-CSx-stage1** |
| **model difference** | **shaPRS-PRSCS** | **r2redux p** | 0.005 | 0.005 |
|  | **shaPRS+LDpred2** | **r2redux p** | 2.992e-04 | 3.067e-4 |
| **LRT of nested vs complex model** | **shaPRS-PRSCS** | **other** | 7.982e-36 | 5.069e-36 |
|  |  | **shaPRS** | 5.136e-13 | 3.451e-13 |
|  | **shaPRS+LDpred2** | **other** | 7.314e-53 | 4.798e-53 |
|  |  | **shaPRS** | 1.251e-17 | 8.696e-18 |

| **Table S5E \| EUR-EAS BRCA- shaPRS comparison against other methods** | | | | |
| --- | --- | --- | --- | --- |
|  |  | **method** | **PRS-CSx** | **PRS-CSx-stage1** |
| **model difference** | **shaPRS-PRSCS** | **r2redux p** | 1.069e-05 | 0.217 |
|  | **shaPRS+LDpred2** | **r2redux p** | 0.001 | 7.501e-10 |
| **LRT of nested vs complex model** | **shaPRS-PRSCS** | **other** | 2.279e-21 | 7.383e-07 |
|  |  | **shaPRS** | 0.619 | 5.113e-15 |
|  | **shaPRS+LDpred2** | **other** | 1.477e-05 | 0.002 |
|  |  | **shaPRS** | 4.290e-28 | 1.003e-53 |

| **Table S5F \| EUR-AFR BMI - shaPRS comparison against other methods** | | | | |
| --- | --- | --- | --- | --- |
|  |  | **method** | **PRS-CSx** | **PRS-CSx-stage1** |
| **model difference** | **shaPRS-PRSCS** | **r2redux p** | 1.590E-01 | 0.00164 |
|  | **shaPRS+LDpred2** | **r2redux p** | 0.915 | 0.0207 |
| **LRT of nested vs complex model** | **shaPRS-PRSCS** | **other** | 7.11E-03 | 1.290E-03 |
|  |  | **shaPRS** | 1.49E-07 | 2.62E-16 |
|  | **shaPRS+LDpred2** | **other** | 2.25E-06 | 8.810E-04 |
|  |  | **shaPRS** | 8.98E-07 | 2.33E-12 |

| **Table S5G \| EUR-AFR height - shaPRS comparison against other methods** | | | | |
| --- | --- | --- | --- | --- |
|  |  | **method** | **PRS-CSx** | **PRS-CSx-stage1** |
| **model difference** | **shaPRS-PRSCS** | **r2redux p** | 9.810E-03 | 0.25 |
|  | **shaPRS+LDpred2** | **r2redux p** | 0.000104 | 0.847 |
| **LRT of nested vs complex model** | **shaPRS-PRSCS** | **other** | 4.16E-09 | 2.32E-06 |
|  |  | **shaPRS** | 2.69E-01 | 8.410E-11 |
|  | **shaPRS+LDpred2** | **other** | 1.47E-12 | 3.810E-07 |
|  |  | **shaPRS** | 9.96E-01 | 7.73E-08 |

| **Table S5H \| EUR-AFR LDL - shaPRS comparison against other methods** | | | | |
| --- | --- | --- | --- | --- |
|  |  | **method** | **PRS-CSx** | **PRS-CSx-stage1** |
| **model difference** | **shaPRS-PRSCS** | **r2redux p** | 1.130E-11 | 0.00494 |
|  | **shaPRS+LDpred2** | **r2redux p** | 9.81E-09 | 0.0475 |
| **LRT of nested vs complex model** | **shaPRS-PRSCS** | **other** | 1.83E-30 | 4.04E-15 |
|  |  | **shaPRS** | 7.94E-01 | 1.170E-03 |
|  | **shaPRS+LDpred2** | **other** | 1.01E-27 | 1.620E-13 |
|  |  | **shaPRS** | 2.97E-01 | 4.11E-05 |

Table of the results for the formal evaluation of model difference between shaPRS and other methods for the cross-ancestry analyses. The **model difference** row shows the p values if there was a difference between shaPRS and the other methods via the ‘r2redux’ r_diff’ and for binary traits, the pROC’ Delong’ tests, respectively. The **LRT of nested vs complex model** row shows the p-values for a likelihood ratio tests that evaluate if adding **shaPRS** or the **other** PRS onto a nested model improves over the complex model of shaPRS+other. For these cross-ancestry analyses shaPRS was evaluated via both PRS-CS (**shaPRS-PRSCS**) and via LDpred2 (**shaPRS+LDpred2**). **A.** EUR-EAS asthma, **B.** EUR-EAS height, **C.** EUR-EAS T2D, **D.** EUR-EAS CAD, **E.** EUR-EAS BRCA, **F.** EUR-AFR BMI, **G.** EUR-AFR height and **H.** EUR-AFR LDL.

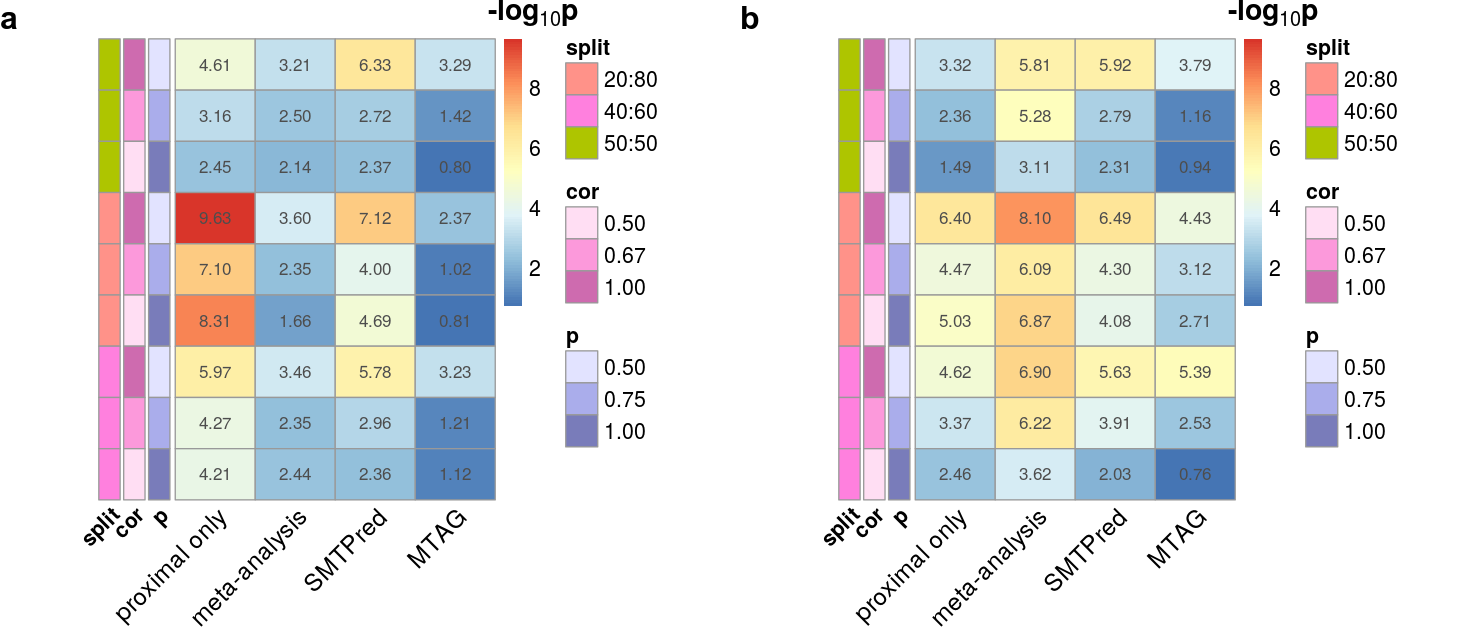

| **Fig S6:** Heatmap of the median ‘r2diff’ -log10(p) model difference between shaPRS and other methods between simulated and predicted phenotypes for selected cross-trait genetic relationships. Warmer colours indicate stronger evidence for a difference between methods. **a.** A genome-wide genetic correlation between proximal and adjunct traits of 0.5 with a heritability of 0.5 from a 1,000 causal variants and no extra heterogeneity created by SNPs of large effect. Sample size N = 14,044, with a proximal/adjunct sample ratio of 50/50, 40/60 or 20/80, and where *cor* is the correlation of effect sizes between SNPs and *P (or causal_S_)* is the fraction of causal SNPs shared between the proximal and adjunct datasets,. *split* is the ratio of the proximal to adjunct dataset sizes. **b.** The same scenario as **a**, with the addition of extra heterogeneity created by five SNPs of large effect that contributed 5% non-shared heritability. Results across the complete set of simulated scenarios are shown in Fig S3. |
| --- |

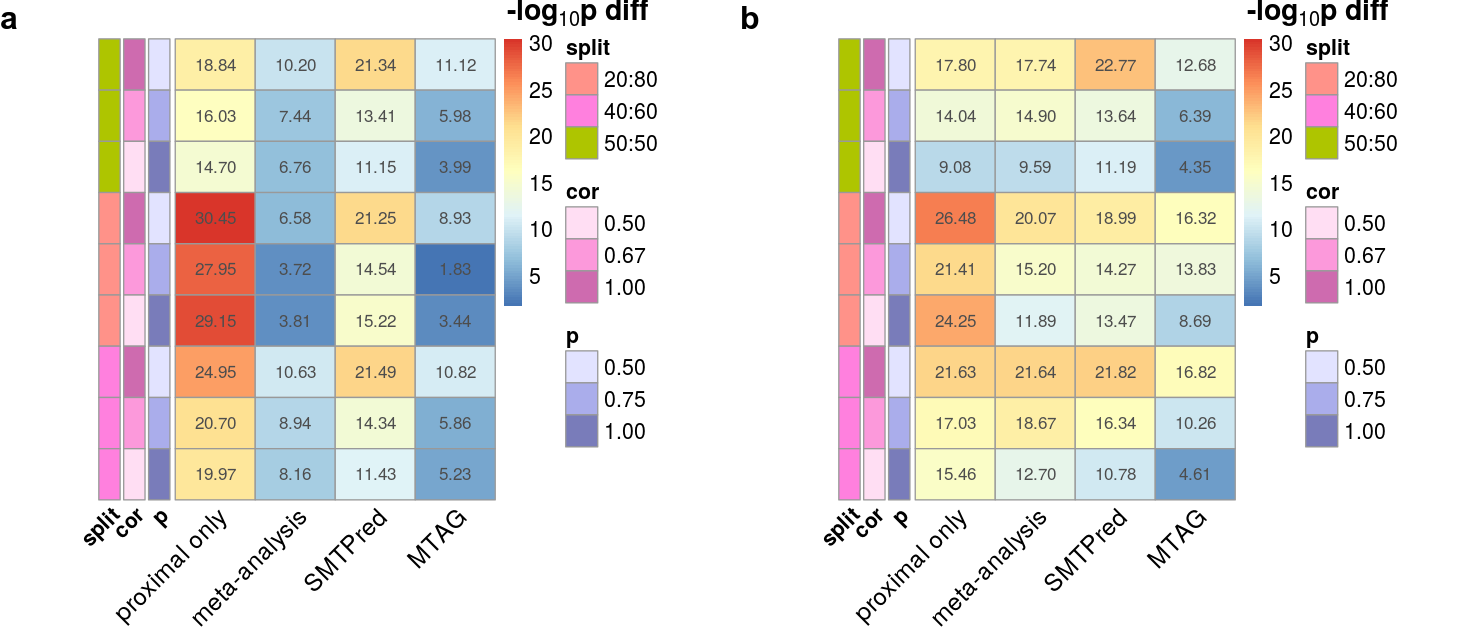

| **Fig S7:** Heatmap of the difference between median -log10(p) values of the likelihood ratio tests of the complex model (which included both shaPRS and the other method) versus the nested model that included either just shaPRS or just the other method for selected cross-trait genetic relationships. Positive values and warmer colours indicate stronger evidence for improving performance by adding shaPRS into the model than adding the other method. **a.** A genome-wide genetic correlation between proximal and adjunct traits of 0.5 with a heritability of 0.5 from a 1,000 causal variants and no extra heterogeneity created by SNPs of large effect. Sample size N = 14,044, with a proximal/adjunct sample ratio of 50/50, 40/60 or 20/80, and where *cor* is the correlation of effect sizes between SNPs and *P (or causal_S_)* is the fraction of causal SNPs shared between the proximal and adjunct datasets,. *split* is the ratio of the proximal to adjunct dataset sizes. **b.** The same scenario as **a**, with the addition of extra heterogeneity created by five SNPs of large effect that contributed 5% non-shared heritability. Results across the complete set of simulated scenarios are shown in Fig S3. |
| --- |
